# A Novel Noise-Resilient and Explainable Machine Learning Framework for Accurate and Robust ECG-Based Heart Disease Diagnosis

**DOI:** 10.1101/2025.07.29.25332377

**Authors:** Abdullah Alhalabi, Saleh Alzahrani, Lamia Al Saikhan, Mahbubunnabi Tamal

## Abstract

An electrocardiogram (ECG) is essential for diagnosing cardiac abnormalities. Automated heartbeat classification enables continuous heart monitoring and early diagnosis. Current machine learning-based methods for heart disease diagnosis from ECG face challenges such as sensitivity to noise, limited adaptivity across different patients, and computational complexity, hindering their adoption. This study introduces a statistical shape model-based method for classifying different heartbeat types: normal, myocardial infarction, premature ventricular contraction, and right bundle branch block. The innovation of this approach lies in its adaptability to shape variability of ECG signals across different patients, as well as its noise-resilience and computational efficiency, allowing for real-time, precise diagnosis of cardiac conditions across diverse ECG morphologies. The model performance was validated on a dataset of 270 patients. It exhibited a strong performance with recall (97.49%), precision (97.73%), F1 Score (97.58%), and accuracy (98.63%) using a support vector machine classifier. When tested with varying levels of signal-to-noise ratio between 12 and −6 dB, the model maintained robust performance with recall (95.49%), precision (95.57%), F1 Score (95.49%), and accuracy (97.44%), demonstrating noise-resilience. Given its high accuracy, robustness, and computational efficiency, this method is well-suited for real-time ECG-based diagnosis and continuous heart monitoring and can be implemented on battery-operated wearables.

## 1. Introduction

Cardiovascular diseases are the leading cause of death worldwide, causing approximately 17.9 million deaths annually, as reported by the World Health Organization (WHO) **[1]**. The electrocardiogram (ECG) is a critical biosignal that captures the electrical activity of the heart muscle using electrodes placed on the body’s skin. It serves as an essential diagnostic tool for evaluating cardiac health and detecting abnormalities in a non-invasive, rapid, painless, and cost-effective manner **[2–4]**. As shown in Figure 1, the ECG signal comprises several key components: the P wave, QRS complex, and T wave, each representing different phases of the cardiac cycle, including the depolarization (contraction) and repolarization (relaxation) of the atria and ventricles.

**Figure 1.**
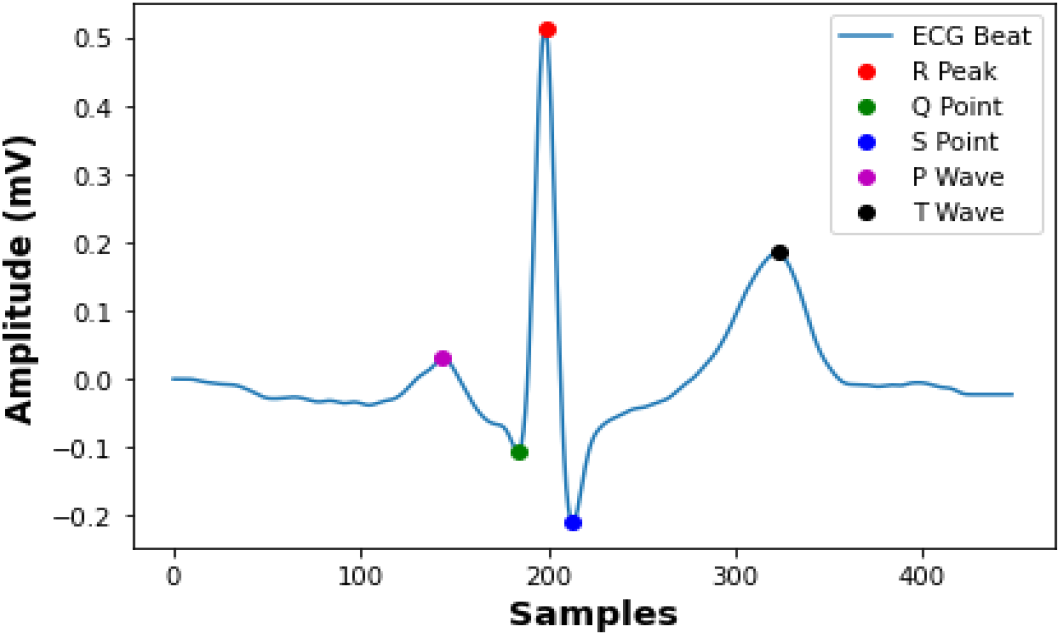
Example of a normal ECG beat with P, Q, R, S, and T points labeled.

With the rise of machine learning (ML) and artificial intelligence (AI) in medical diagnostics, automated ECG-based analysis has gained significant traction due to its ability to provide accurate and fast interpretations **[5]**. Advanced ML and AI models can detect subtle patterns and anomalies in ECG data that might be overlooked by human observers, leading to improved accuracy. Additionally, it can analyze large amounts of ECG data much faster than humans, enabling prompt diagnosis and timely intervention, which is crucial in acute cardiac events. The automation of ECG interpretation can also reduce medical care costs and allow healthcare professionals to focus on more complex cases and direct patient care.

Recently, automated ECG classification has become a growing focus in research. The study in **[6]** proposed a deep learning approach for classifying ECG beats into normal and abnormal using the MIT-BIH Arrhythmia Database **[7]**. They developed a deep neural network (DNN) by conducting various trials and manually configured network parameters, including the number of hidden layers, the number of neurons in each layer, the activation function, and the number of learning steps. The best configuration they found includes seven hidden layers, with the number neurons making up each layer as follows: 5, 10, 30, 50, 30, 10, and 5, respectively Although the model achieved an accuracy of over 99%, it was limited to differentiating between normal and abnormal beats without identifying the types of abnormalities.

The study in **[8]** introduces a method for classifying heartbeats based on features derived from time-frequency analysis of ECG. Using the MIT-BIH Arrhythmia Database, the study classifies heartbeats into 5 categories. Features extracted include RR interval, higher-order statistical (HOS) measures like skewness and kurtosis, and pseudo-energy features from Wigner-Ville Distribution (WVD) across nine time-frequency windows. An ensemble of decision trees is employed for classification. The proposed method achieves high overall sensitivity and positive predictivity of 99.67% and 98.92%, respectively. However, a drawback of this method is the high computational cost, limiting its applicability for online heartbeat classification with limited hardware environments.

In **[9]**, the authors introduce a real-time, patient-specific method using 1D convolutional neural networks (CNNs) to classify cardiac arrhythmias. The authors validated the system’s performance using the MIT-BIH Arrhythmia Database and demonstrated that it yields a better classification performance than other state-of-the-art methods for detecting ventricular and supraventricular ectopic beats. However, this method is limited in that it cannot be applied for different patients as the model should be trained specifically for each individual patient, which is time- and labor-consuming.

The study in **[10]** introduces the optimum-path forest (OPF) classifier as a novel approach for heartbeat classification. The classifier’s performance was studied against support vector machine (SVM), Bayesian, and multilayer perceptron (MLP) classifiers across six feature extraction methods for arrhythmia analysis in ECG. The authors used the MIT-BIH Arrhythmia Database and classified heartbeats into five categories: normal (N), supraventricular ectopic beat (S), ventricular ectopic beat (V), fusion beat (F), and unknown beat (Q). In average, the OPF outperforms MLP and SVM in classification time and accuracy and provides comparable performance to the Bayesian classifier. However, this method requires extensive preprocessing of data for optimal performance.

The study in **[11]** presents an eleven-layer deep convolutional neural network (CNN) to classify ECG signals into four categories as follows: normal (N), atrial fibrillation (A-Fib), atrial flutter (AFL), and ventricular fibrillation (V-Fib). Three databases were used: used the MIT-BIH Arrhythmia Database, the MIT-BIH Atrial Fibrillation Database **[12]**, and Creighton University Ventricular Tachyarrhythmia Database **[13]**. The model achieved 94.90% accuracy, 99.13% sensitivity, and 81.44% specificity. The drawbacks of this model are that it takes a long training time and needs a large amount of data (big data) for training.

The study in **[14]** introduces a knowledge-based approach for automatic heartbeat classification. The method extracts several morphology- and rhythm-based features, followed by applying QRS clustering to minimize the impact of potential interpretation errors. Finally, each cluster is labeled using a rule-based classifier. The method has been validated on the MIT-BIH Arrhythmia Database and achieved superior classification performance compared to other state-of-the-art methods for identifying ventricular and supraventricular ectopic beats. However, this method is unsuitable for real-time applications.

The study in **[15]** explores various machine learning algorithms for detecting and classifying cardiac arrhythmias into 14 different classes using the UCI Arrhythmia Dataset **[16]**. The study uses two feature extraction techniques and compares multiple models such as Naive Bayes, SVM, Random Forests, and Neural Networks. The study found that a combined approach of SVM and RF classifiers outperformed single classifiers and improved accuracy to 77.4%. However, this accuracy is relatively modest and highlights the need for further improvement in arrhythmia classification models.

Table 1 below presents a summary of the primary limitations of the previous methods. Another issue is that most of the methods in the literature were tested on relatively high quality or preprocessed ECG records, but their performance often deteriorates when tested on noise-corrupted signals. In real-world scenarios, noise is an inevitable factor, particularly in ECGs collected via wearable devices over extended periods. Moreover, numerous studies employ the intra-patient paradigm for model evaluation, where ECG data from the same patients are included in both training and testing sets **[17]**. This practice leads to overfitting and an overestimation of model performance. In contrast, the inter-patient paradigm divides the training and test sets on a subject-by-subject basis, which means the model is trained on ECG data from one group of patients and tested on data from a completely different group. This approach is considered more suitable for the development of models intended for clinical use **[18]**, as it better reflects real-world scenarios where models must generalize to unseen patients. The review in **[17]** compares 11 studies that examined both intra-patient and inter-patient paradigms. The average performance reveals notable differences in key metrics. For intra-patient classification, the overall accuracy is 98.39%, F1-score is 95.52%, sensitivity is 93.51%, positive predictive value is 92.78% and specificity is 99.19%. In contrast, for inter-patient classification, the accuracy drops to 90.15%, F1-score to 83.89%, sensitivity to 78.16%, positive predictive value to 62.82%, and specificity to 93.86%. This decline in performance is attributed to morphological variations in ECGs among patients, despite having the same abnormalities **[19]**.

**Table 1.**
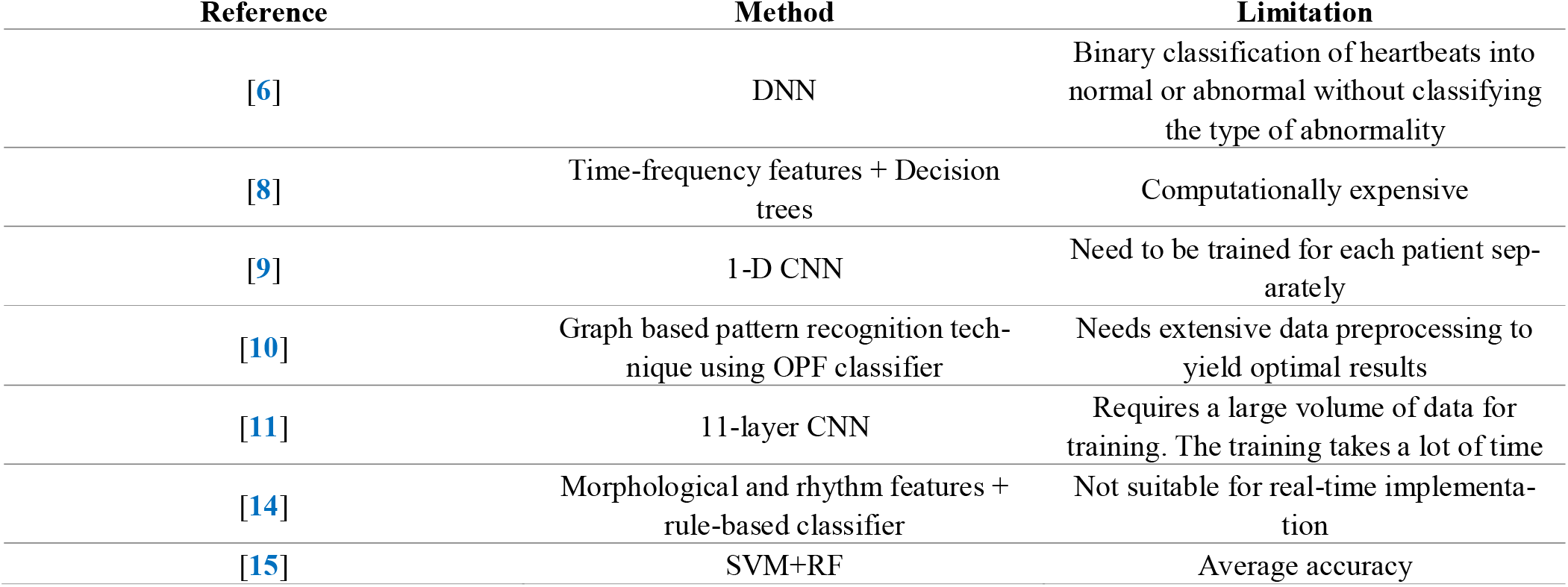
Limitations of previous studies on ECG classification.

Based on the preceding review of literature, there is a clear need for a new classification method that offers higher adaptability to ECG shape of different patients, works in real-time with minimal computational demands, and more accurately classifies heartbeats under noisy scenarios. In response, this thesis proposes a novel method based on SSMs, which is designed to meet the above criteria. To the best of our knowledge, SSMs have not previously been applied for heartbeat classification in ECG signals. The remainder of this paper is organized as follows: In Section **2**, we explain how the proposed algorithm works and its validation process. In Section **3**, we present the results and discuss the main findings of this study. Finally, in Section **4**, we conclude this paper and provide recommendations for future work.

## 2. Materials and Methods

### 2.1. Statistical Shape Models (SSMs)

SSMs are mathematical models that represent the statistical distribution of shapes and are used in various fields such as medical imaging and computer vision **[20, 21]**. These models are built from sets of shapes by capturing their variations using statistical techniques like principal component analysis (PCA) **[22]**. PCA serves to simplify complex, high-dimensional data while retaining as much of the original variability as possible. It identifies the key directions in which the data varies (principal components). By projecting the data onto these principal components, PCA effectively reduces the number of dimensions and helps to represent the original data with a smaller set of parameters **[22]**. SSMs are used in many applications including image segmentation, shape analysis, and shape reconstruction, where they help accurately delineate anatomical structures, understand morphological variations, and support tasks such as disease diagnosis, product design, and therapy planning **[23]**.

SSMs are derived from a training set of shapes. Every shape in this set is characterized by a group of n landmarks in d-dimensional space, which serve as correspondence points, indicating specific locations on the shape. To represent the shape, all the node coordinates are combined into a single vector as shown in Equations 1 and 2 below for 2D and 3D shapes **[24]**, respectively, where ***T*** denotes transpose.

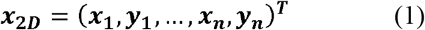

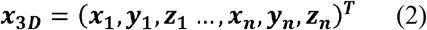

here, ***x***_**2*D***_ and ***x***_**3*D***_ are **2n × 1** and **3n × 1** vectors, respectively. The shapes in the training set need to be aligned into to a common coordinate system using Procrustes alignment **[24]** to remove effects due to translation, rotation, and scaling. This alignment ensures that the variability captured by PCA is due to shape differences rather than differences in rotation, translation, and scaling. Once the shapes are aligned, PCA is applied to capture the variability within the data, which allows for the synthesis of new shapes similar to those in the original training set. To do this, we first compute the mean shape 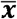 by averaging the aligned shapes as follows **[24]**:

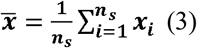

where ***n***_***s***_ is the number of training samples. Next, we compute the normalized covariance matrix of the data as follows **[24]**:

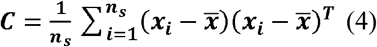

After this, eigenvalue ***(λ***_***i***_) and the corresponding eigenvector ***(p***_***i***_ ) can be obtained from the covariance matrix C by solving Eq. (5) below **[24]**.

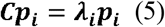

The eigenvalues are sorted in descending order by default. Each eigenvector corresponds to a direction of variation, and the corresponding eigenvalue indicates the magnitude of the variation in that direction. In other words, the eigenvectors correspond to the modes of variation, and the eigenvalues represent the variance along each mode. The aligned shapes in the initial set can be then approximated using a linear combination of these modes as follows **[22, 24]**:

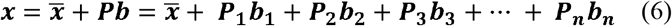

Typically, we want to capture a sufficient amount of the total variation in the data, for example 95% **[24]**, therefore we select the first K eigenvectors (principal components) that yields the required variation, which can be calculated from the following equation **[24]**:

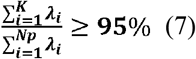

where ***Np*** is the total number of eigenvalues (or principal components) calculated from the covariance matrix, and ***λ***_***i***_ is the *i*^*th*^ eigenvalue obtained from the covariance matrix. The adjustment of b in Eq. (6) above allows for the generation of new shapes within the range of variability captured by the PCA by deforming the mean shape 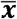. Vector b is expressed as **[22]**:

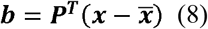

The variance of each parameter ***b***_***i***_ is controlled by the corresponding eigenvalue ***λ***_***i***_. To ensure that new generated examples have a shape similar to the ones in the training dataset, the shape parameter b is usually restricted within 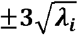 **[22]**, where ***λ***_***i***_ is the eigenvalue corresponding to the *i*^*th*^ mode of variation.

### 2.2. Heartbeat Data Preparation

This study focuses on constructing a SSM for both normal and abnormal heartbeats, specifically targeting the following types: (i) normal (N), (ii) myocardial infarction (MI), (iii) premature ventricular contraction (PVC), and (iv) right bundle branch block (RBBB). Normal ECG beats exhibit a regular pattern with distinct P, QRS, and T waves. The QRS complex is narrow (under 120 ms), indicating normal ventricular depolarization, and the overall rhythm is regular with uniform R-R intervals. PVC is an arrhythmia characterized by extra heartbeats that originate from the ventricles, causing premature and abnormal beats **[25]**. While often benign and asymptomatic, frequent PVCs can lead to palpitations or indicate underlying heart issues. PVC beats are characterized by an early and wide QRS complex (greater than 120 ms), typically without a preceding P wave **[25]**. The T wave often points in the opposite direction of the QRS complex, and the beat occurs prematurely, interrupting the normal rhythm. MI occurs when blood flow to part of the heart is blocked, causing heart muscle damage due to oxygen deprivation. It often presents with chest pain, shortness of breath, and ECG changes like ST-segment elevation **[26]**, which vary with the stage of infraction and is more prominent in specific ECG leads according to the location of the infarction within the heart. MI is a medical emergency that can lead to severe complications if untreated. RBBB is a cardiac conduction abnormality seen in ECG where the electrical impulse is delayed or blocked in the right bundle branch of the heart’s conduction system. This results in a characteristic widening of the QRS complex (greater than 120 ms) with distinctive patterns in lead V1, often showing a double-peak or notched pattern **[27]**. RBBB can be caused by conditions like right cardiomyopathy, pulmonary embolism, or it may occur in otherwise healthy individuals without apparent heart disease **[27]**. It is important for clinicians to assess its underlying cause when detected in an ECG.

A total of 900 ECG records, each sampled at 500 Hz, were utilized in this study. The durations of these records range between 5 and 60 seconds. The ECG data was sourced from the PhysioNet/Computing in Cardiology Challenge 2021 **[28]**, available on Physionet **[29]**. It comprises a diverse collection of databases representing a wide range of demographic groups. Specifically, the data used in this thesis were obtained from the following databases under the PhysioNet/Computing in Cardiology Challenge 2021: i) The China Physiological Signal Challenge 2018 (CPSC2018) Database **[30]**, including the unused data from CPSC 2018 (the CPSC-Extra Database) - China, ii) The Physikalisch Technische Bundesanstalt (PTB-XL) Database **[31]** - Germany iii) The Georgia 12-lead ECG Challenge (G12EC) Database **[32]** - USA, iv) The Shaoxing and Ningbo Hospital ECG Database **[33–35]** – China. Normal and PVC beats were extracted from lead II, RBBB beats from V1, and MI beats were extracted from multiple leads as follows: II, III, aVL, aVF, V5, and V6. The ECG recordings were initially processed using two 4th-order Butterworth filters. The first one is a high-pass filter with cutoff frequency of 0.67 Hz to remove baseline wander. The second one is a 40Hz low-pass cutoff frequency that was used to reduce noise. Subsequently, ECG beats were segmented by first detecting the R peaks and identifying the midpoint of the R-R interval. Visual inspection was done to assess segmentation accuracy, and manual adjustments were made when needed by selecting a specific number of samples on either side of this midpoint between consecutive R peaks to capture a complete heartbeat with all key waveform components (P, QRS, and T). An enhanced version of Elsahmrany’s algorithm **[36]** was utilized for R peak detection, although it has not been published **[37]**. Existing segmentation algorithms are not perfect when it comes to splitting ECGs into heartbeat with all key waveform components. We performed the segmentation manually to ensure that our shape model is built correctly. Therefore, our work focuses on the classification part, assuming that the segmentation was already done correctly. In each record, the first and last beats, along with any distorted beats, were excluded. Figure 2 below shows examples of heartbeats from different classes, highlighting the shape variability between ECG beats obtained from different patients.

**Figure 2.**
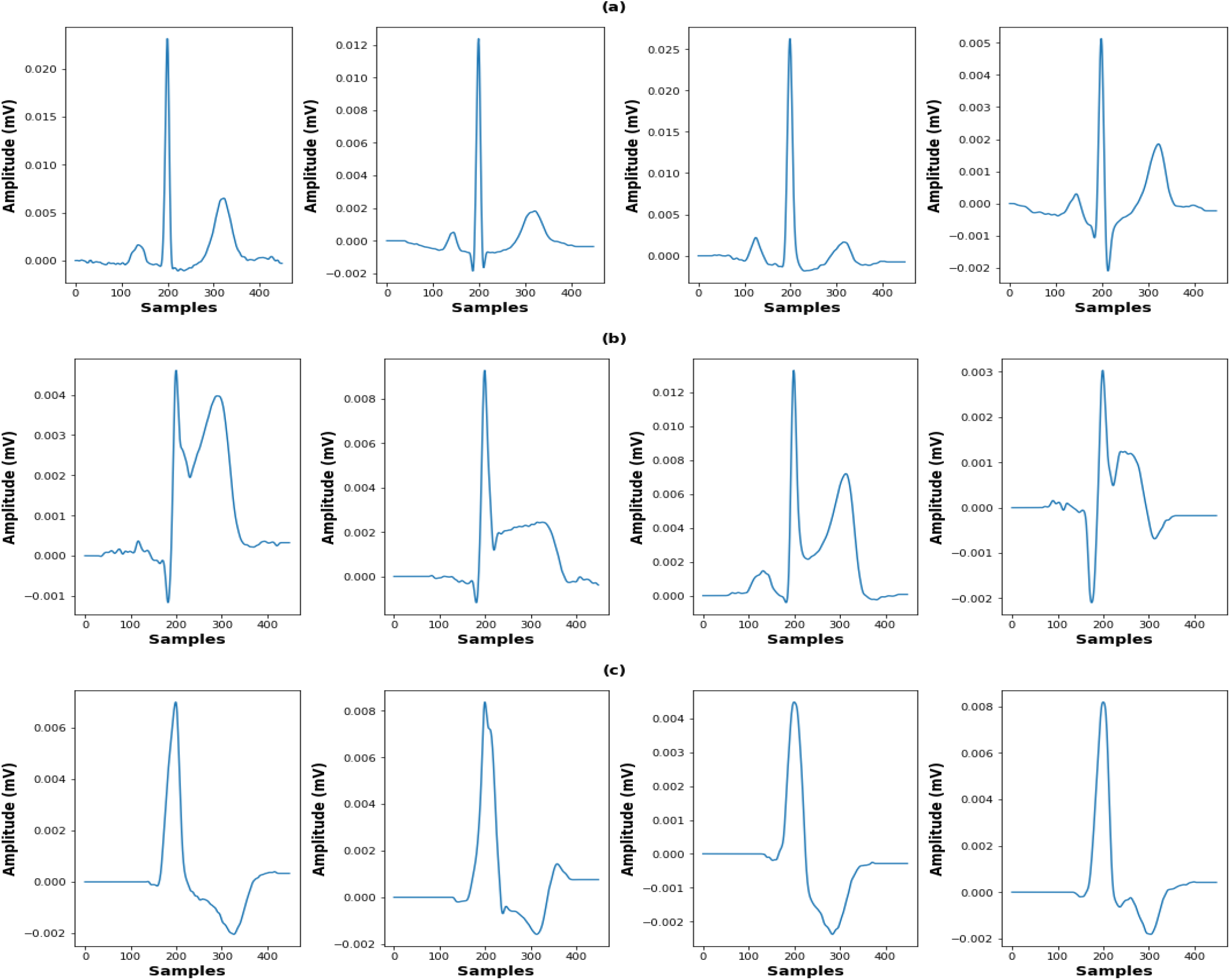
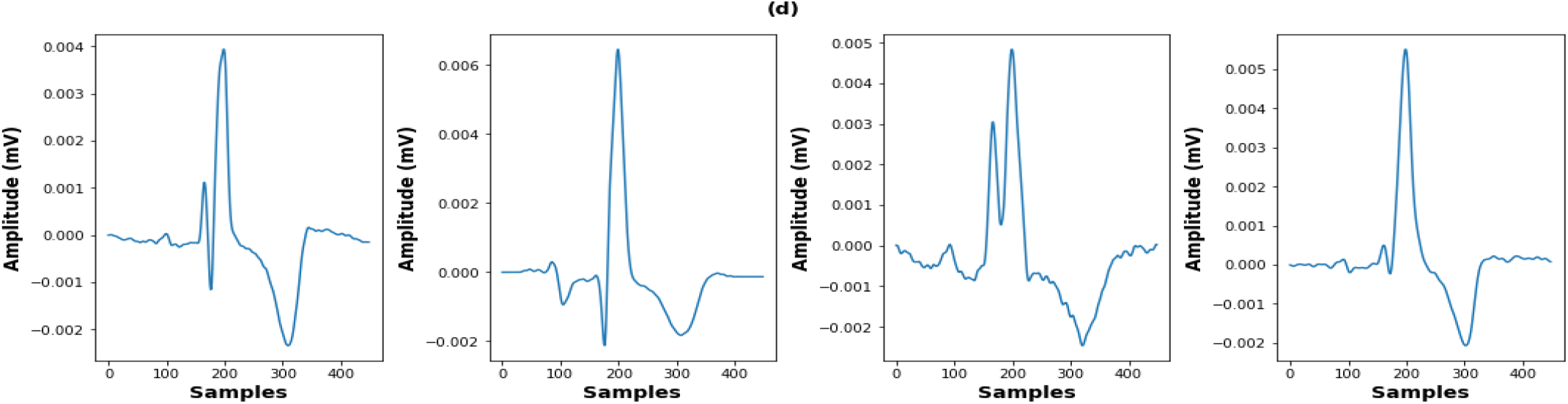
Examples of heartbeats for different types (a) Normal (N), (b) Premature Ventricular Contraction (PVC), (c) Myocardial Infarction (MI), and (d) Right Bundle Branch Block (RBBB). Normal beats have a sharp, narrow QRS complex with regular shape. MI beats show an elevated ST segment. PVC beats exhibit broad QRS complexes with inverted T waves. RBBB beats are characterized by widened QRS with a distinctive double-peak or notched pattern. This figure highlights variations in heartbeat morphologies within the same class.

After segmentation, heartbeats were aligned using the R peaks as reference points, with the start of each beat padded with its initial value, applying a variable number of samples for each beat to ensure alignment with a predefined R peak position. Similarly, the end of each beat was padded with its final value using a variable number of samples for each beat to achieve the predefined number of samples for equalization. The total number of samples per beat was then standardized to 450 by trimming excess samples both the start and end. Next, each beat was vertically shifted so that its minimum value was set to zero. Finally, each beat underwent area under the curve (AUC) normalization by dividing each sample within a beat by the sum of all the samples in that beat, resulting in an AUC equal to one. Finally, to center each beat’s baseline around zero, all beats were shifted such that the first sample of each beat was set to zero. Figures 3 and 4 below present the aligned beats for the four classes along with the mean beat ± two times standard deviation that reflects the shape variations among different beats.

**Figure 3.**
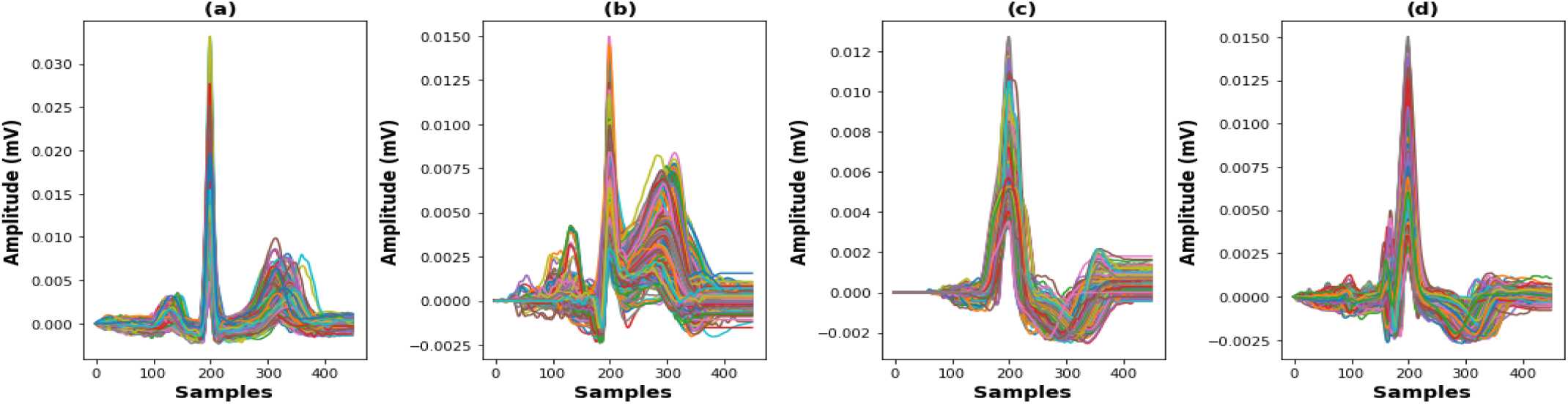
Aligned and normalized beats from the 630-patient training set, showing (a) N, (b) MI, (c) PVC, and (d) RBBB.

**Figure 4.**
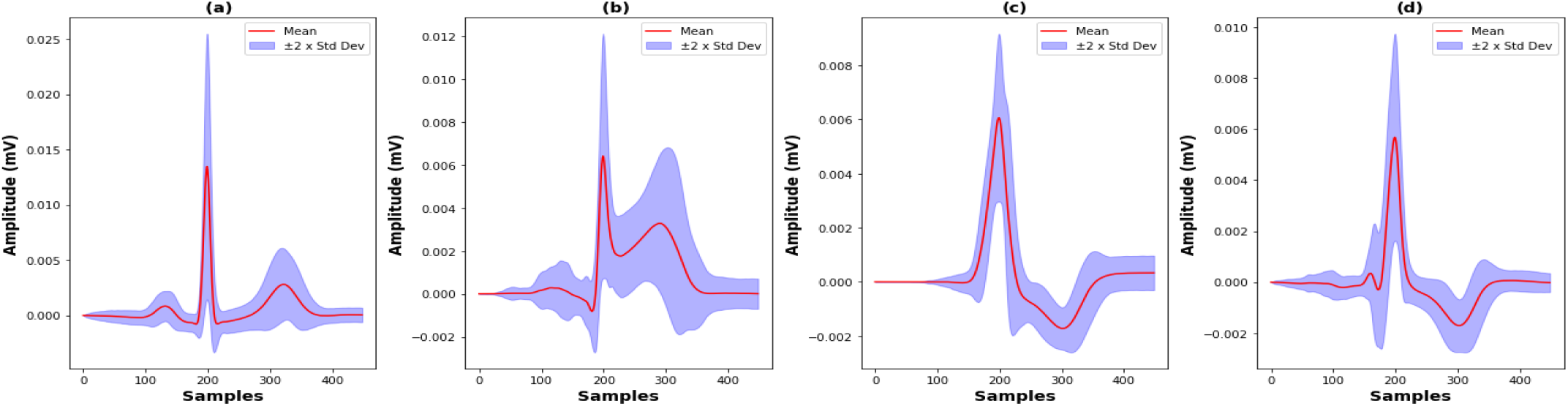
Plot of four heartbeat classes as mean ± 2 × the standard deviation for each class (a) N, (b) MI, (c) PVC, and (d) RBBB.

The 900 ECG records were divided, with 70% used for building the shape model and training machine learning classifiers, and the remaining 30% were used for performance testing. Table 2 below summarizes the number of patients and heartbeats per class. For Normal and RBBB classes, we downsampled the number of beats for every patient in order to maintain data balance and prevent model bias toward majority classes, since the PVC and MI classes contained a smaller number of beats compared to Normal and RBBB. This disparity arises due to the fewer MI patients included in this study and the fact that patients with PVCs exhibited only a few PVC beats, with the majority being non-PVC. In contrast, for Normal and RBBB patients, all ECG records exhibited only Normal or RBBB beats, allowing us to take a representative subset. Therefore, it is unnecessary to include all Normal and RBBB beats in each record as all of them were found to be almost identical and that is why we took only a subset of beats from each patient to prevent data imbalance.

**Table 2.** Description of the number of patients and heartbeats per class.

| **Shape Model Building and Classifier Training** |  |  |  |  |  |
| --- | --- | --- | --- | --- | --- |
|  | **N** | **MI** | **PVC** | **RBBB** | **Total** |
| **Number of Patients** | 225 | 42 | 174 | 189 | 630 |
| **Number of Heartbeats** | 521 | 410 | 718 | 653 | 2302 |
| **Classifier Performance Testing** |  |  |  |  |  |
|  | **N** | **MI** | **PVC** | **RBBB** | **Total** |
| **Number of Patients** | 108 | 18 | 63 | 81 | 270 |
| **Number of Heartbeats** | 264 | 222 | 345 | 300 | 1131 |

### 2.3. Shape Model Fitting for Heartbeat Classification

To perform fitting, we first construct a unified shape model for all heartbeat types by aligning them all together based on R peak positions and applying PCA to extract the eigenvectors (principal components) and their corresponding eigenvalues. The reconstruction of a target beat is expressed as a linear combination of a mean beat and a set of principal components derived from PCA. The goal is to find the optimal ***b*** values, which are the coefficients that scale the contribution of each principal component to the final reconstruction. Mathematically, the target beat is approximated as the mean beat plus the sum of the principal components, each multiplied by its corresponding ***b*** value.

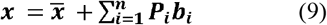

here, ***x*** represents the beat that we are trying to reconstruct or fit, while 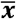 is the mean beat. The principal components capture the key variation patterns from the data, and the ***b*** values control how much of each component is used to reconstruct the beat. Solving for ***b*** values involves minimizing the difference between the actual target beat and the reconstructed beat, which can be done using least-squares. Least-squares fitting is a mathematical approach used to find the best-fit line or model for a given set of data by minimizing the sum of the squared differences between the observed data points and the predicted values generated by the model. The linear model can be expressed as:

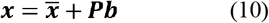

where ***x*** is the actual target beat which we are trying to approximate, 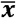is the mean beat, and ***P*** is the matrix of principal components. Finally, ***b*** is the vector of coefficients that indicates how much each principal component contributes to the reconstructed beat. The goal of least-squares fitting is to minimize the sum of the squared residuals (difference between the actual target beat and the reconstructed beat). The equation for the residual is:

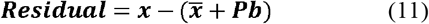

where 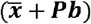 is the fitted beat based on the coefficients ***b***. The objective is to find the values of ***b*** that minimize the residual (the difference) between the actual target beat ***x*** and the reconstructed beat 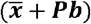 as follows:

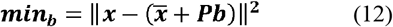

here, 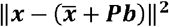 represents the sum of the squared residuals. To find the best set of ***b*** values, we solve the least-squares problem in Equation 12. This equation gives the optimal values for the ***b***, which minimize the squared error between the reconstructed beat 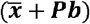 and the actual target beat ***x***. To solve this, we first expand the squared norm and set it to zero:

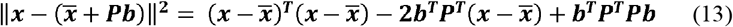

Next, we take the derivative of this expression with respect to ***bT*** and set it to zero:

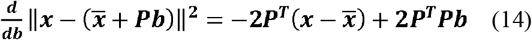

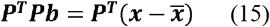

To isolate ***b***, we multiply both sides by the inverse of ***P***^***T***^***P*** to get:

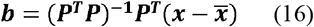

Since the principal components are orthogonal (a property inherent to PCA), the matrix ***P***^***T***^***P*** is the identity matrix. This simplifies the solution to:

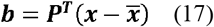

Which is identical to Equation 8 presented earlier. This approach ensures that the beat is reconstructed using the selected principal components and their corresponding coefficients. These coefficients are determined by multiplying the transpose of the principal components’ matrix ***P*** with the difference between the target beat and the mean beat.

Via visual inspection, we found that the use of 19 principal components (resulting in 19 ***b*** values) yields a very good fit across all classes. These 19 principal components account for 99.55% of total data variability. Although increasing the number of components beyond 19 improves fitting quality, employing more components than necessary can lead to greater model deformation, causing it to fit noise. This reduces the model’s resilience to noise and may compromise classification accuracy. Thus, determining an adequate number of components is essential to balance between fitting quality and noise-resilience. Figure 5 displays examples of fitted heartbeats for the four classes. For heartbeat classification, the combination of values (19 ***b*** values) that were used to fit each beat serve as features for training a machine learning algorithm. To classify a new beat, we fit it using our shape model and feed the resulting values into the trained classifier to determine the type of that beat.

**Figure 5.**
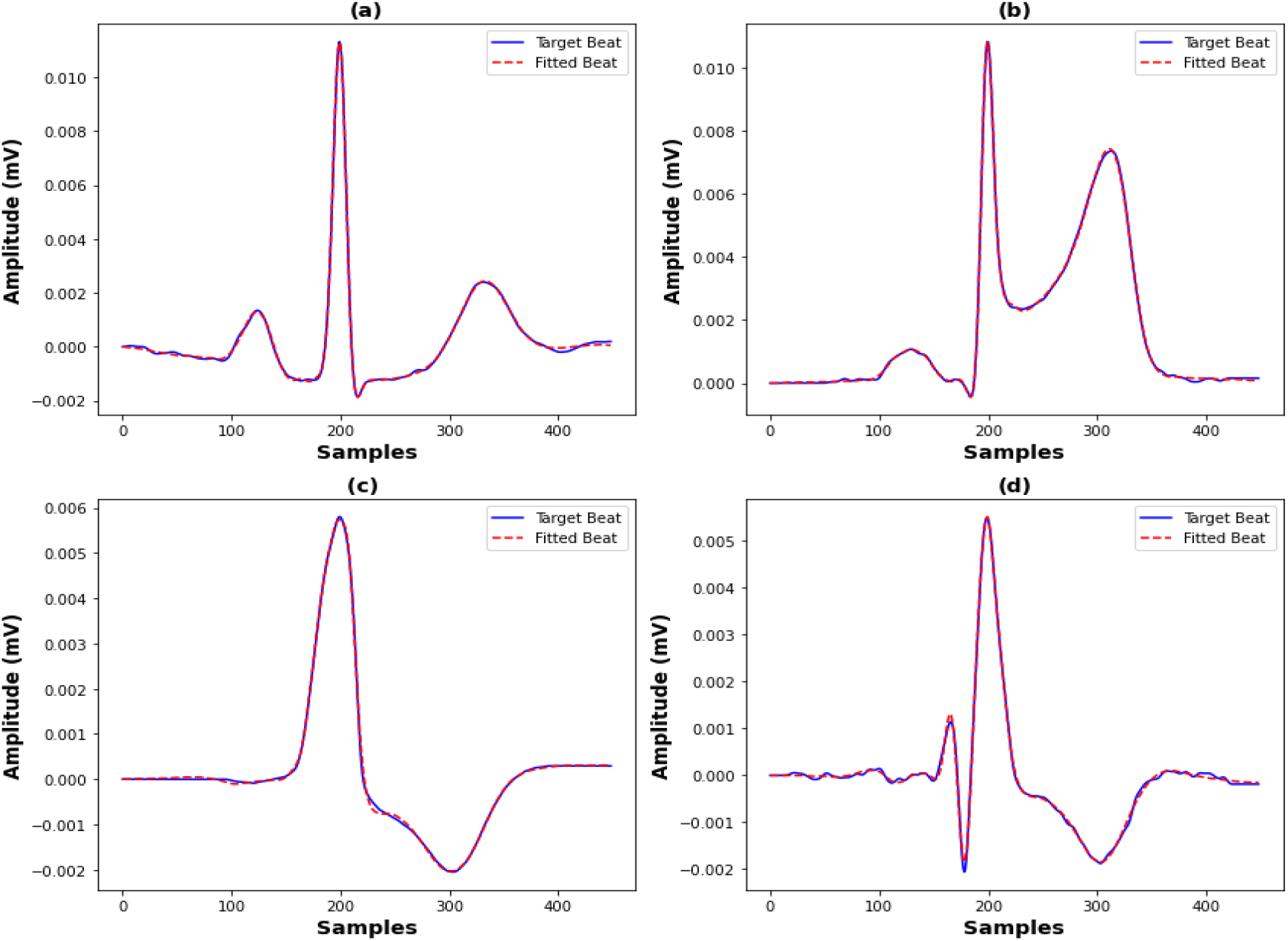
Shape model fitting of each beat type using 19 principal components (a) N, (b) MI, (c) PVC, and (d) RBBB.

### 2.4. Evaluation of The Proposed Method

The proposed model was implemented in *Spyder* (*Python* 3.10.9) on a laptop with a 1.8 GHz 8-Core Intel Core i7 processor, 8 GB of RAM, running Windows 10. Heartbeat classification performance was assessed using four metrics: recall, precision, F1-score, and accuracy (Equations 18–21). The recall metric is used to measure how well a model can correctly identify true positive cases without missing them. It is computed by calculating the proportion of true positives out of all actual positive cases. High recall is crucial since failing to detect an abnormality result in a missed diagnosis. On the other hand, precision focuses on the accuracy of the positive predictions made by the model. It measures the proportion of true positive results out of all predicted positive cases. High precision means that the model is usually correct when it predicts a positive case. High precision is crucial in situations where false positives may result in significant consequences, such as a wrong diagnosis. The F1-score is the harmonic mean of both recall and precision and is useful to account for both false positives and false negatives. It offers a balanced view of model performance where neither precision nor recall can be prioritized over the other. Finally, accuracy is a metric that measures the proportion of all correct predictions (both true positives and true negatives) out of the total number of predictions made. It is useful for getting a general idea about a model performance across all classes. By considering all these four metrics, we can draw a more complete picture of model performance and understand its strengths and weaknesses.

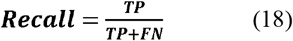

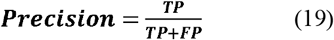

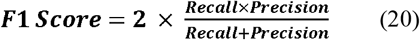

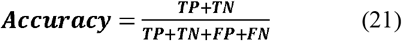

here, true positives (TP) refer to cases where the model correctly classifies a heartbeat into its actual class. A false negative (FN) happens when the model fails to recognize a heartbeat’s correct class. False positives (FP) occur when the model incorrectly predicts a heartbeat as belonging to a class that it does not actually belong to. A true negative (TN) refers to cases where the model accurately excludes a class, correctly identifying a heartbeat as not belonging to a particular class.

Four different classifiers were used in this study, which are random forest (RF), support vector machine (SVM), extra trees (ET), and quadratic discriminant analysis (QDA), form *Scikit-Learn* package **[38]** with default settings as detailed in Appendix A (Table A1). We also tested our method’s robustness under noisy ECG data. Gaussian noise was added at different signal-to-noise ratios (SNRs): 12 dB, 6 dB, 0 dB, and −6 dB. The noise was added according to Equation 22 below, were 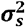 is the variance of the ECG signal and 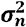 is the variance of noise. The noise hinders the accurate identification of fiducial points in ECG, such as onset, offset and peak location of ECG waves. Therefore, our method is expected to perform well in the presence of noise since it depends on the overall shape rather than specific points as well as it does not fit to noise. Figure 6 illustrates this concept by displaying an example of RBBB beat fitting at varying noise levels.

**Figure 6.**
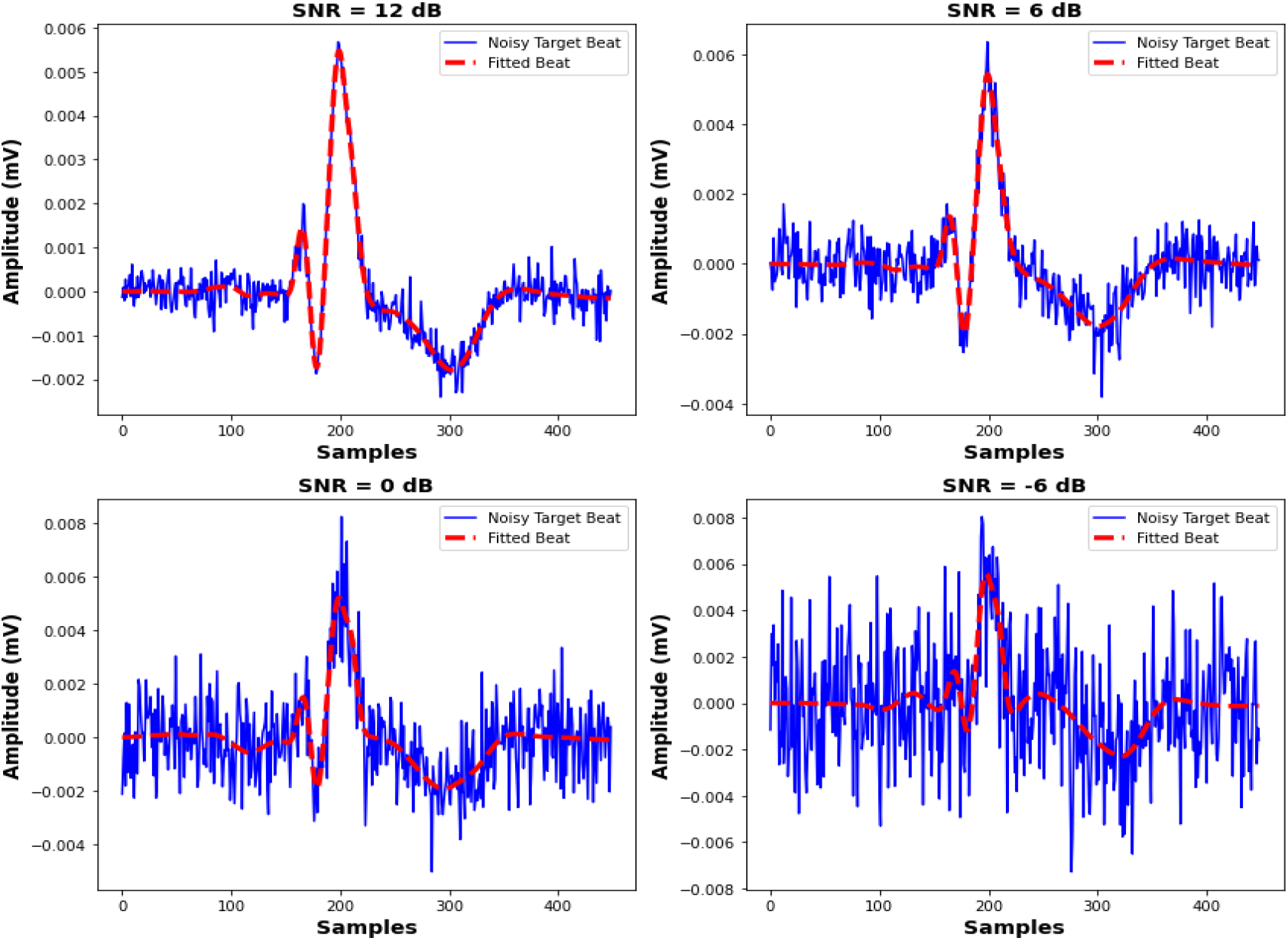
Shape model fitting for an RBBB beat at various SNR levels (12 to −6 dB).

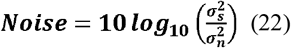

## 3. Results and Discussions

### 3.1 Heartbeat classification in noise-free signals

The confusion matrices of classification are shown in Figure 7 for all four classifiers. Additionally, Table 3 lists the evaluation metrics for them. The RF classifier achieved perfect recall (100.00%) for the Normal class, which means that all Normal beats were correctly identified, as can be seen in the confusion matrix. MI recall is slightly lower at 97.30% as five MI beats were misclassified as Normal and one as RBBB. PVC recall is high at 99.13% with only three PVC beats being misclassified as RBBB. On the other hand, RBBB has the lowest recall at 95.67%, due to the highest number of misclassifications into other classes. There are eleven RBBB beats that were classified mistakenly as PVC and two as Normal. Precision is perfect for MI (100.00%), which means that there are no other beats that were wrongly classified as MI. The precision is high for RBBB (98.63%) since only three PVC beats and one MI beat were misclassified as RBBB. In contrast, the precision for Normal and PVC was lower due to the higher number of misclassifications. F1-scores across classes remain above 97% and overall accuracy above 98%, indicating excellent overall performance. In summary, the RF model demonstrates strong performance, achieving an average accuracy of 99.03% in heartbeat classification.

**Table 3.**
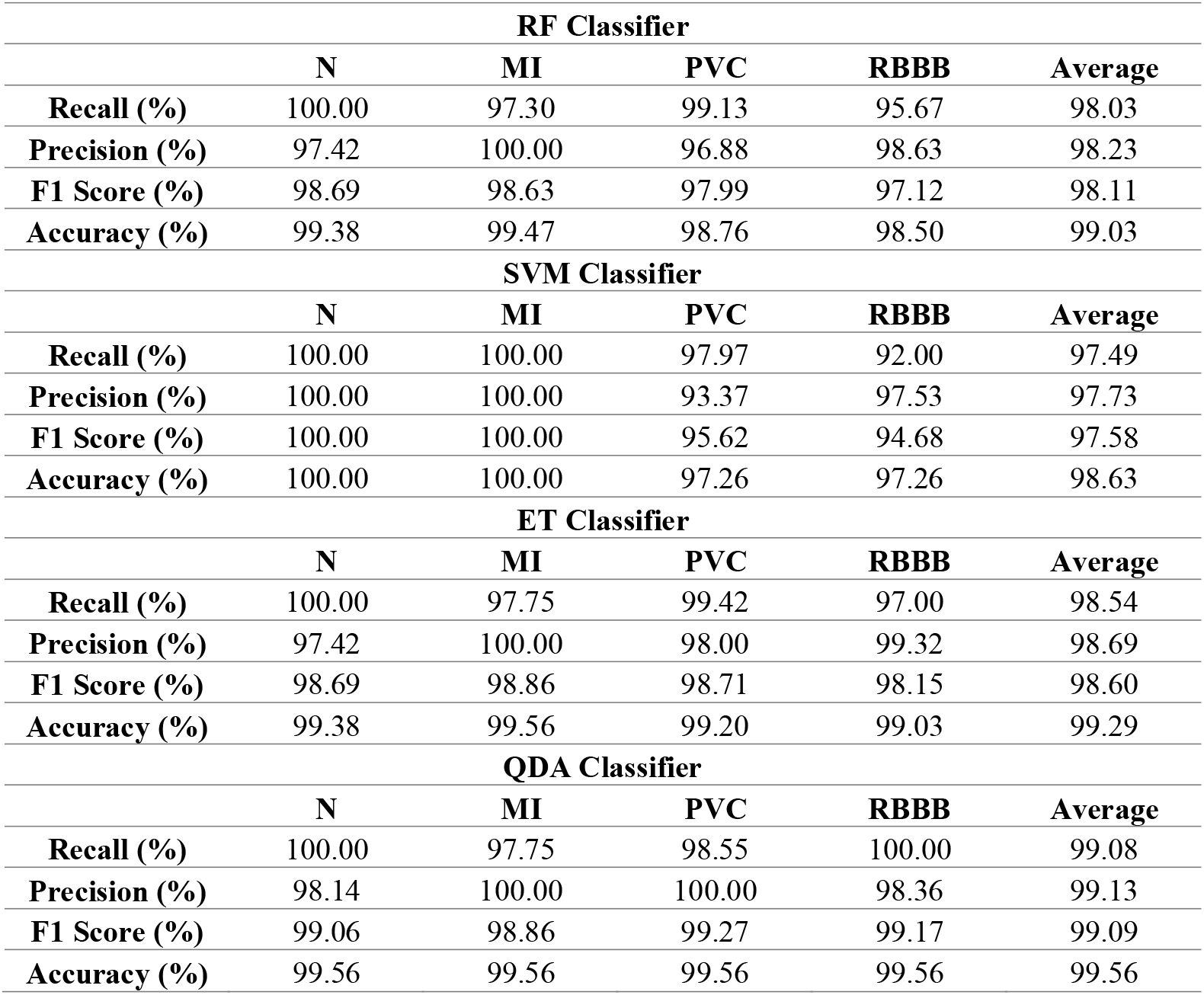
Evaluation metrics for heartbeat classification using different classifiers.

**Figure 7.**
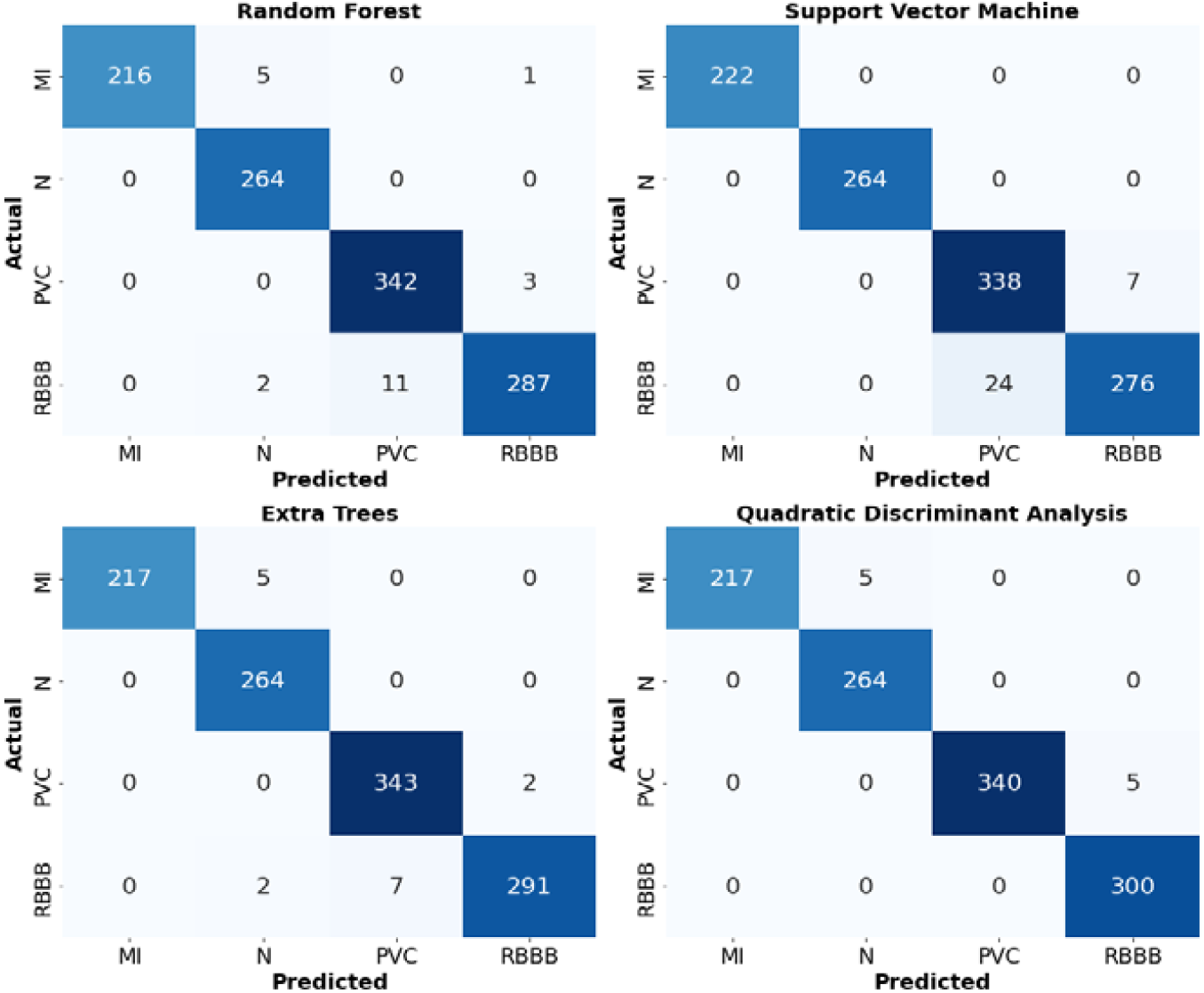
Confusion matrices for heartbeat classification across four machine learning classifiers: RF, SVM, ET, and QDA. Each matrix displays the predicted versus actual counts for four heartbeat types: N, MI, PVC, and RBBB.

The SVM classifier achieved 100% recall and precision for both the Normal and MI classes. This means that all beats belonging to these classes were correctly identified by the model, and no beats form other classes were misclassified to be belonging to these classes. However, recall and precision for PVC drop to 97.97% and 93.37%, respectively, and for RBBB they decrease to 92.00% and 97.53%, respectively. This reflects a larger confusion between these two classes as compared to the RF model. There were twenty-four RBBB beats misclassified as PVC, and seven PVC beats were identified wrongly as RBBB. The F1-scores follow a similar trend, with lower values for PVC (95.62%) and RBBB (94.68%). Despite these challenges, the SVM model still achieves a high overall accuracy of 98.63%.

The ET model has similar recall and precision for Normal and MI class as the RF classifier. It only predicted five MI beats wrongly as Normal and two RBBB beats as Normal, while RF misclassified five MI as Normal, two RBBB beats as Normal, and one MI beat as RBBB. In addition, the ET classifier exhibits larger recall and precision for RBBB and PVC classes compared to the RF. This suggests that it has a better ability to distinguish between PVC and RBBB beats, though some misclassification persists. The F1-scores across classes are similarly high, with the lowest being 98.15% for RBBB heartbeats. The model’s accuracy for each class is excellent, exceeding 99%. The average accuracy of 99.29% across all heartbeat classes highlights the strong overall performance of this classifier.

The QDA model misclassifies five MI beats as Normal, as observed in the RF and ET classifiers. However, it shows superior performance when classifying PVC and RBBB beats, with a misclassification of only five PVC beats as RBBB. Recall is 100.00% for both Normal and RBBB classes, and slightly lower for MI (97.75%) and PVC (98.55%). Precision is also high across all classes, with the lowest value being 98.14% for Normal and a perfect 100.00% for MI and PVC. The F1-scores mirror the other metrics, with the lowest being 98.86% for MI and the highest being 99.27% for PVC. With an overall accuracy of 99.56%, the QDA classifier stands out for its strong performance across all metrics and classes.

In summary, this comparison highlights the strengths and weaknesses of each classifier. All classifiers showed robust overall performance in classifying different heartbeat types. It was noticed that Normal and MI beats were well classified in general, though slight misclassifications were observed in RF, ET, and QDA classifiers. On the other hand, the SVM perfectly classified all Normal and MI beats with no errors. The most variation occurs in the classification of PVC and RBBB beats, especially in the SVM classifier which exhibited weaker performance compared to the other classifiers. The QDA model excelled at handling PVC and RBBB classification with only minor errors. Overall, the QDA classifier demonstrated the best performance among other classifiers.

### 3.2. Heartbeat Classification Under Noisy Conditions

The proposed method was evaluated across a range of SNRs (from 12 dB to −6 dB) to assess its robustness under different noise conditions. Table 4 below lists the evaluation indices for each classifier across all SNRs.

**Table 4.** Comparison of different classifiers under various SNRs.

| **RF Classifier** |  |  |  |  |  |
| --- | --- | --- | --- | --- | --- |
|  | **12 dB** | **6 dB** | **0 dB** | **-6 dB** | **Average** |
| **Recall (%)** | 98.14 | 96.81 | 92.00 | 82.02 | 92.24 |
| **Precision (%)** | 98.28 | 96.83 | 92.41 | 85.10 | 93.16 |
| **F1 Score (%)** | 98.20 | 96.80 | 91.77 | 80.56 | 91.83 |
| **Accuracy (%)** | 99.07 | 98.28 | 95.45 | 89.57 | 95.59 |
| **SVM Classifier** |  |  |  |  |  |
|  | **12 dB** | **6 dB** | **0 dB** | **-6 dB** | **Average** |
| **Recall (%)** | 97.42 | 97.30 | 96.49 | 90.75 | 95.49 |
| **Precision (%)** | 97.63 | 97.42 | 96.50 | 90.74 | 95.57 |
| **F1 Score (%)** | 97.50 | 97.35 | 96.49 | 90.62 | 95.49 |
| **Accuracy (%)** | 98.59 | 98.50 | 98.01 | 94.65 | 97.44 |
| **ET Classifier** |  |  |  |  |  |
|  | **12 dB** | **6 dB** | **0 dB** | **-6 dB** | **Average** |
| **Recall (%)** | 98.64 | 97.29 | 91.76 | 81.70 | 92.35 |
| **Precision (%)** | 98.76 | 97.30 | 92.30 | 85.96 | 93.58 |
| **F1 Score (%)** | 98.69 | 97.28 | 91.39 | 79.94 | 91.83 |
| **Accuracy (%)** | 99.34 | 98.54 | 95.18 | 89.48 | 95.64 |

| QDA Classifier |  |  |  |  |  |
| --- | --- | --- | --- | --- | --- |
|  | 12 dB | 6 dB | 0 dB | -6 dB | Average |
| **Recall (%)** | 92.66 | 81.72 | 72.68 | 54.60 | 75.42 |
| **Precision (%)** | 93.58 | 87.80 | 83.48 | 71.71 | 84.14 |
| **F1 Score (%)** | 92.21 | 78.95 | 65.66 | 46.86 | 70.92 |
| **Accuracy (%)** | 95.67 | 89.12 | 84.17 | 75.95 | 86.23 |

The provided confusion matrices and line graph in Figures 8 and 9 display the classification performance of the RF classifier across various noise levels. At 12 and 6 dB, the RF classifier demonstrates strong performance, with only a few misclassifications. However, as the noise level increases, classification errors start to escalate notably, particularly between PVC and RBBB which are very similar classes. At 0 dB, the recall and precision fall to 92.00% and 92.41%, respectively, and they decline further to 82.02%, and precision drops to 85.10% at the lowest SNR of −6 dB. The line graph of recall, precision, F1 score, and accuracy reveals a gradual decline in the evaluation metrics as the noise level increases. Overall, the RF classifier appears to maintain a good performance till 0 dB, with all metrics exceeding 90%. These results indicate that the RF classifier has a certain degree of resilience to noise, but higher noise levels (−6 dB) significantly impact its overall classification accuracy.

**Figure 8.**
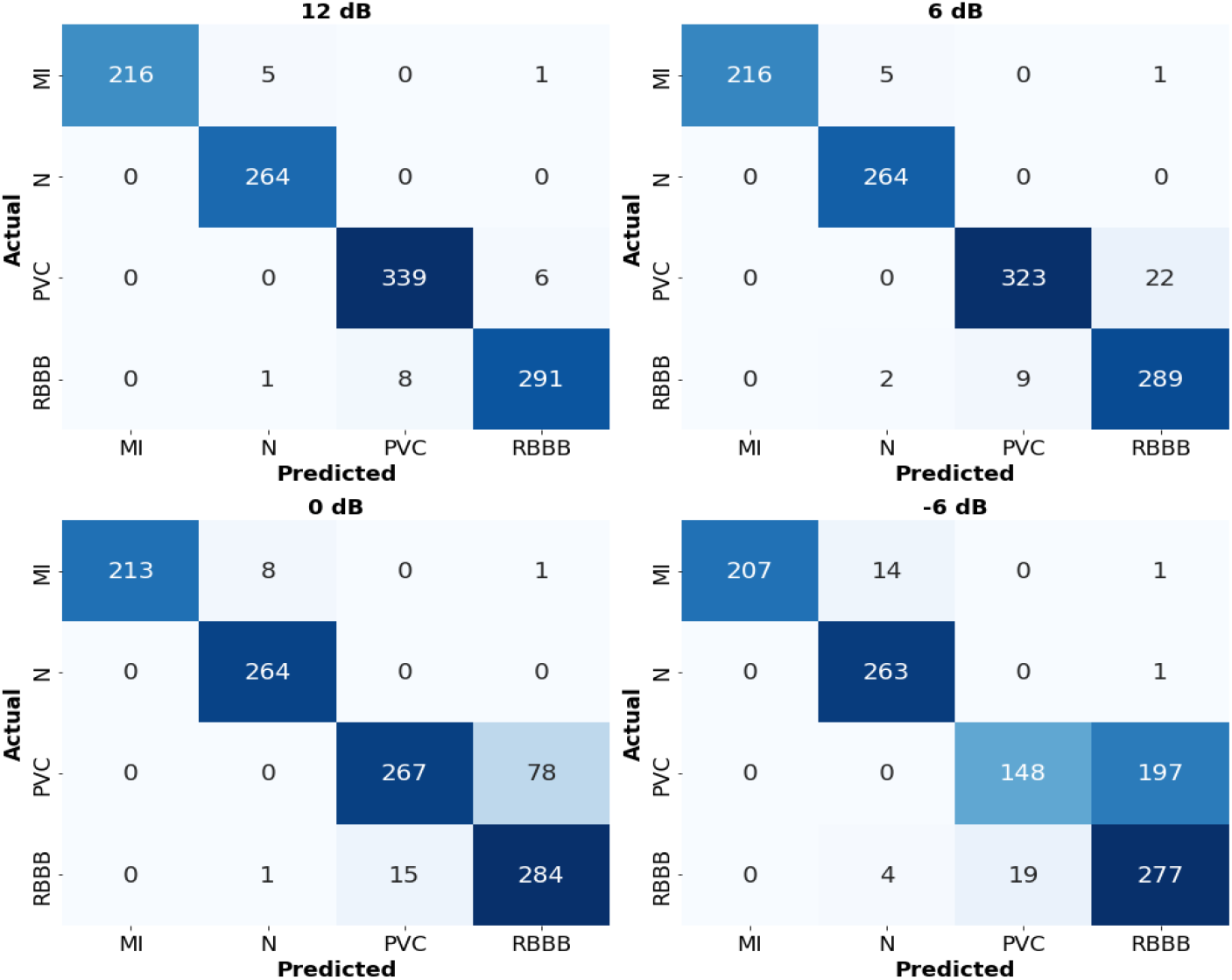
Confusion matrices of heartbeat classification using RF classifier with different SNR levels.

**Figure 9.**
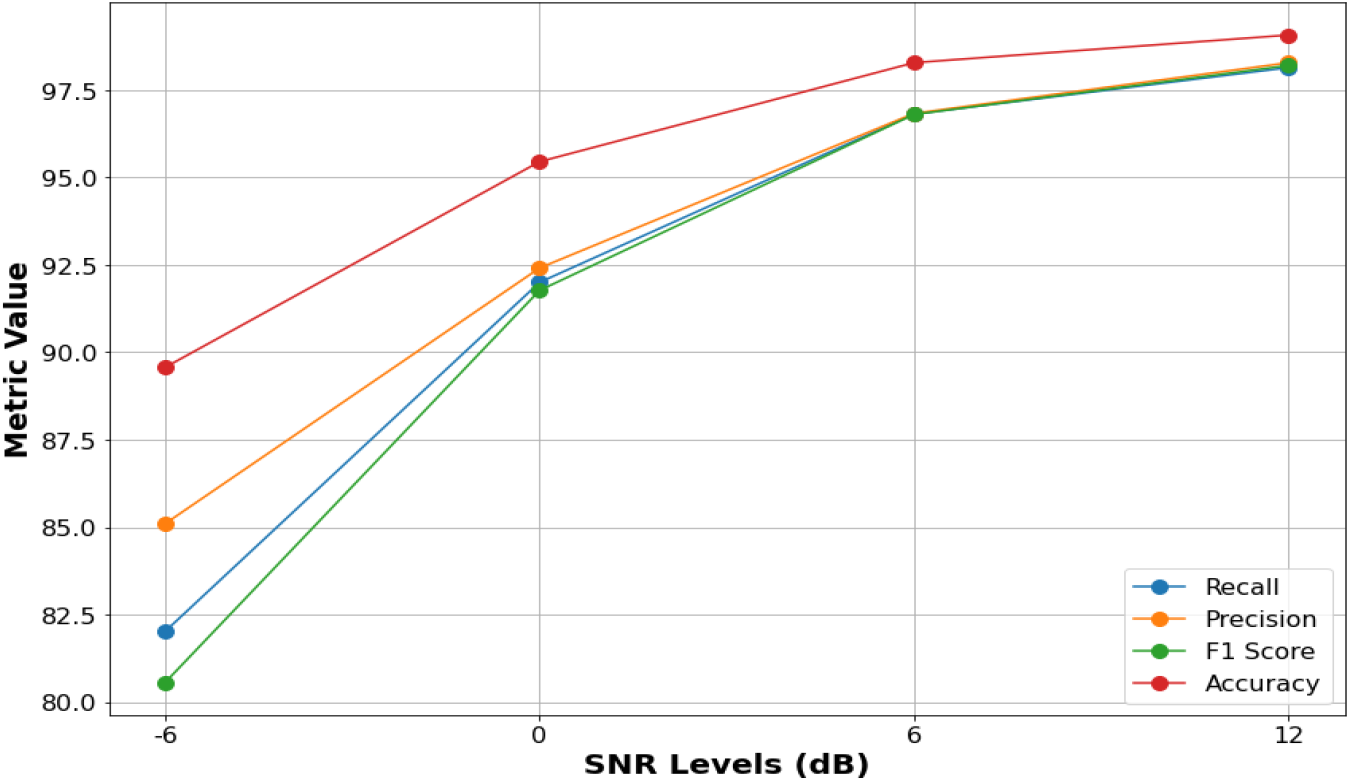
Recall, precision, F1-score, and accuracy of RF classifier with different SNR levels.

The provided confusion matrices and performance metrics in Figures 10 and 11 illustrate the behavior of the SVM classifier in heartbeat classification across different amounts of noise. At 12 dB and 6 dB, the SVM classifier achieves strong classification performance, with high recall and precision, exceeding 97%. As noise increases to 0 dB, performance remains high with only a minor decline. At the lowest SNR (−6 dB), classification errors increase between PVC and RBBB beats, with recall and precision declining to 90.75% and 90.74%, respectively. Notably, the classifier identified all N and MI beats correctly across all SNRs, except only one Normal beat as RBBB at −6dB. Overall, the SVM model shows strong noise-resilience and noticeably outperforms the RF classifier at high noise levels (0 dB and −6 dB).

**Figure 10.**
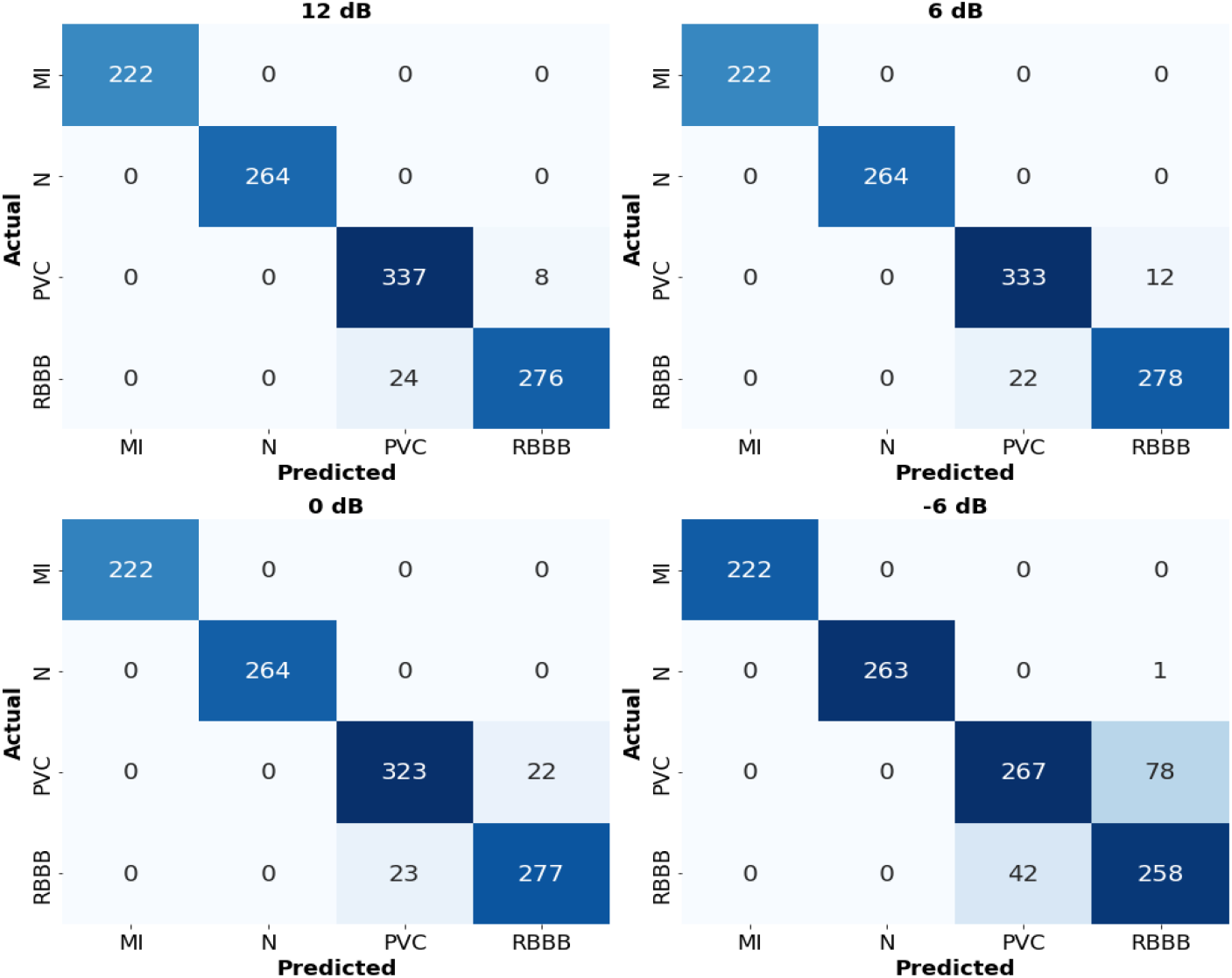
Confusion matrices of heartbeat classification using SVM classifier with different SNR levels.

**Figure 11.**
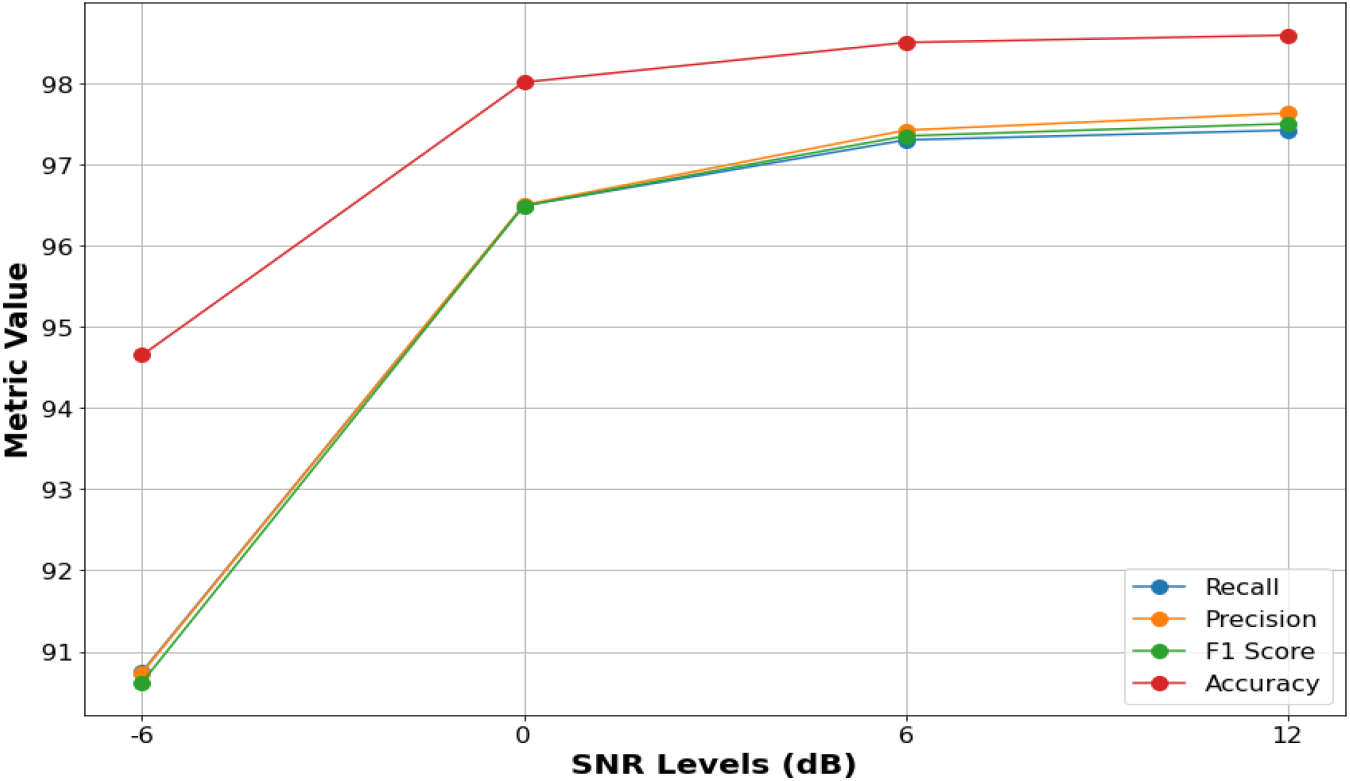
Recall, precision, F1-score, and accuracy of SVM classifier with different SNR levels.

The provided confusion matrices and line graph in Figures 12 and 13 illustrate the classification performance of the ET classifier at various noise levels. At 12 dB, the classifier shows a high F1 score (98.69%), along with minimal errors across all classes. As the SNR decreases to 6 dB, some misclassifications begin to appear, particularly between PVC and RBBB, though F1 score remains high at 97.28%. By 0 dB, performance degradation becomes more evident, with F1 score dropping to 91.39%. At −6 dB, F1 score declines significantly to 79.94%, with a notable increase in misclassifications among similar classes like PVC and RBBB and some overlap between MI and normal beats. The line graph of recall, precision, F1 score, and accuracy shows a consistent decline across all metrics as noise increases, highlighting reduced classifier reliability in noisy conditions. These results indicate that while the ET classifier performs well in low-noise environments, its performance deteriorates considerably under high noise levels.

**Figure 12.**
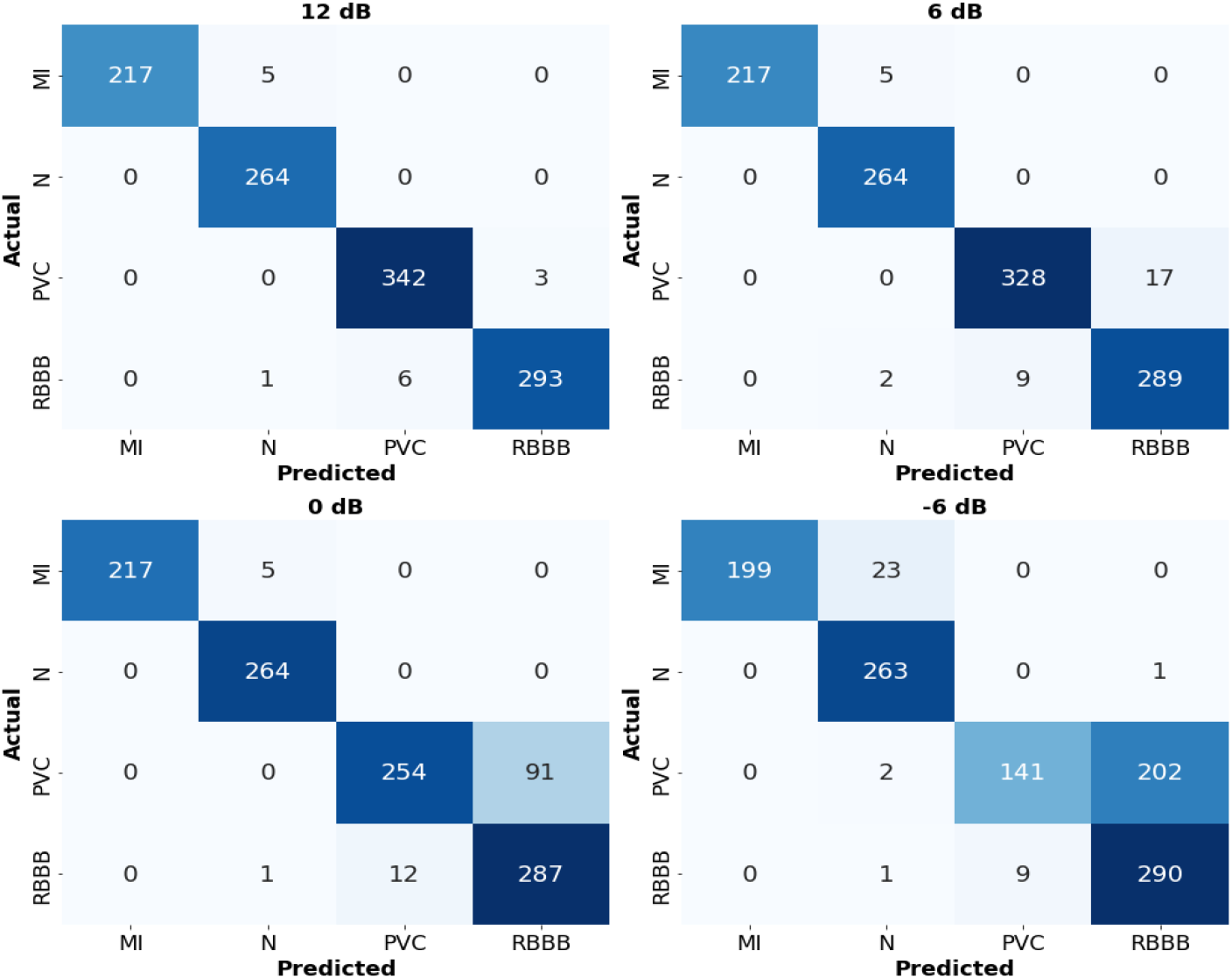
Confusion matrices of heartbeat classification using ET classifier with different SNR levels (a) 12 dB, (b) 6 dB, (c) 0 dB, and (d) −6 dB.

**Figure 13.**
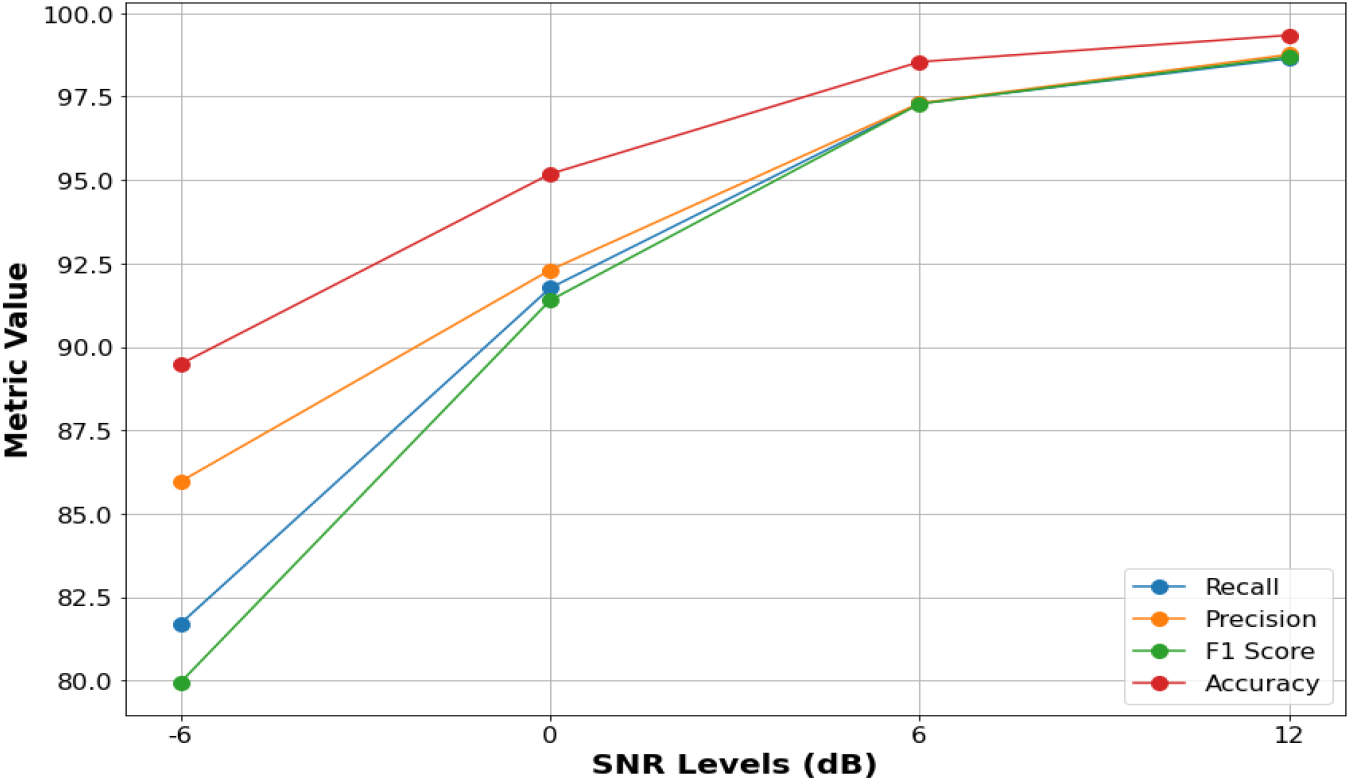
Recall, precision, F1-score, and accuracy of ET classifier with different SNR levels.

The provided confusion matrices and line graph in Figures 14 and 15 illustrate the impact of noise on heartbeat classification using QDA. As the SNR decreases, there is a noticeable degradation in the model’s ability to accurately classify beats. At 12 dB, QDA shows average performance, with an F1 score of 92.21%. As the SNR drops to 6 dB and subsequently to 0 dB, the number of misclassifications rises significantly, particularly between similar classes (PVC and RBBB), and F1 score drops to 78.95% and further to 65.66% at 0 dB. This trend continues at −6 dB, where QDA performs the worst, with F1 score declining sharply to 46.68% and substantial confusion among different heartbeat types is present. Compared to other classifiers, QDA is more sensitive to noise and its performance deteriorates faster than that of SVM, RF, and ET as noise levels increase. The line graph of performance metrics shows a considerable decline across all metrics, especially F1 score and recall, which highlights the QDA model’s difficulty in correctly identifying heartbeat types under noisy conditions. Overall, this comparison has shown that the SVM outperforms all other classifiers in terms of overall robustness to noise as it preserves higher accuracy and balanced performance across metrics in noisy conditions. RF classifier follows with a good performance as well. The ET comes next, while the QDA was found to be the most susceptible to noise as its performance metrics deteriorated significantly as noise levels increased.

**Figure 14.**
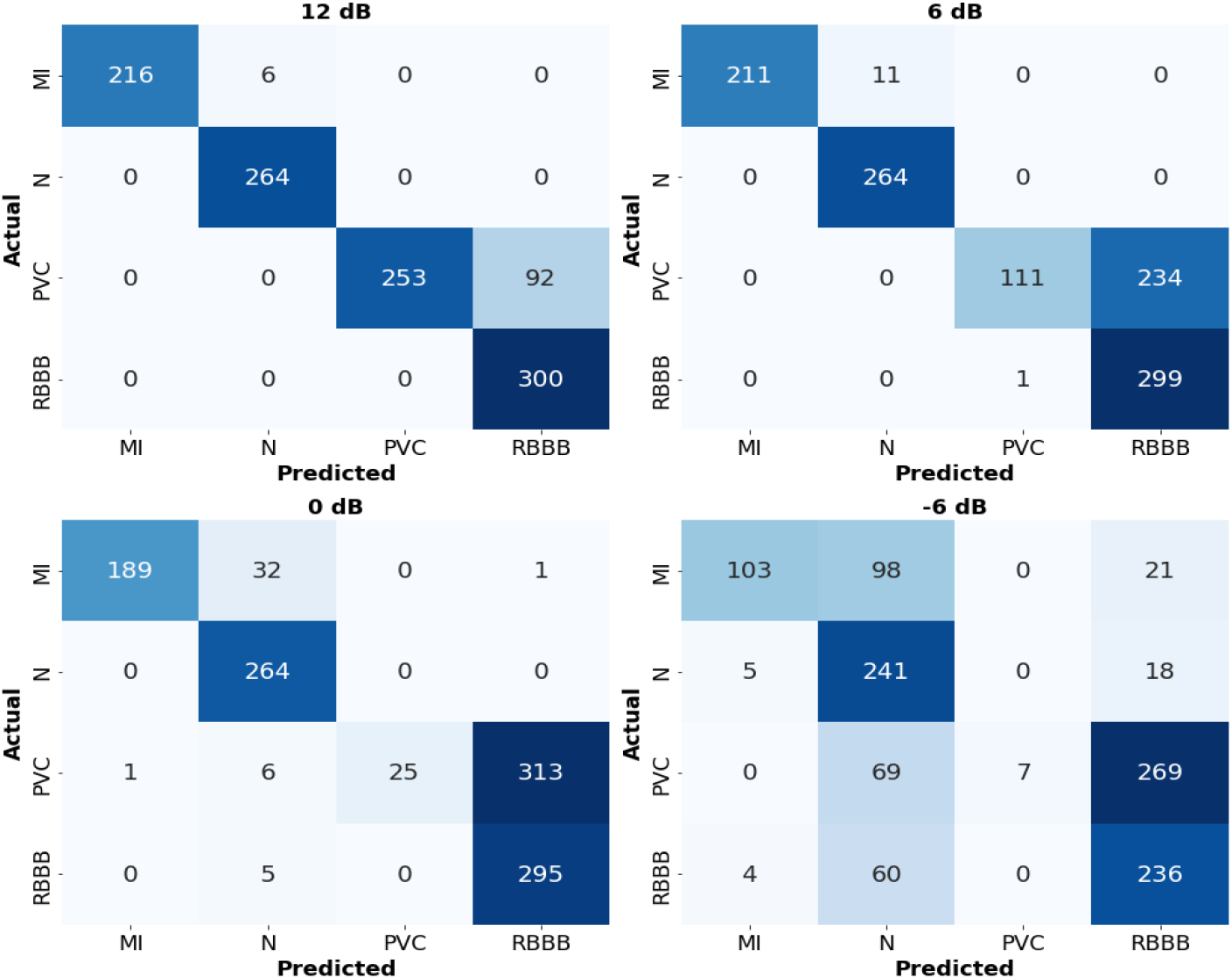
Confusion matrices of heartbeat classification using QDA classifier with different SNR levels.

**Figure 15.**
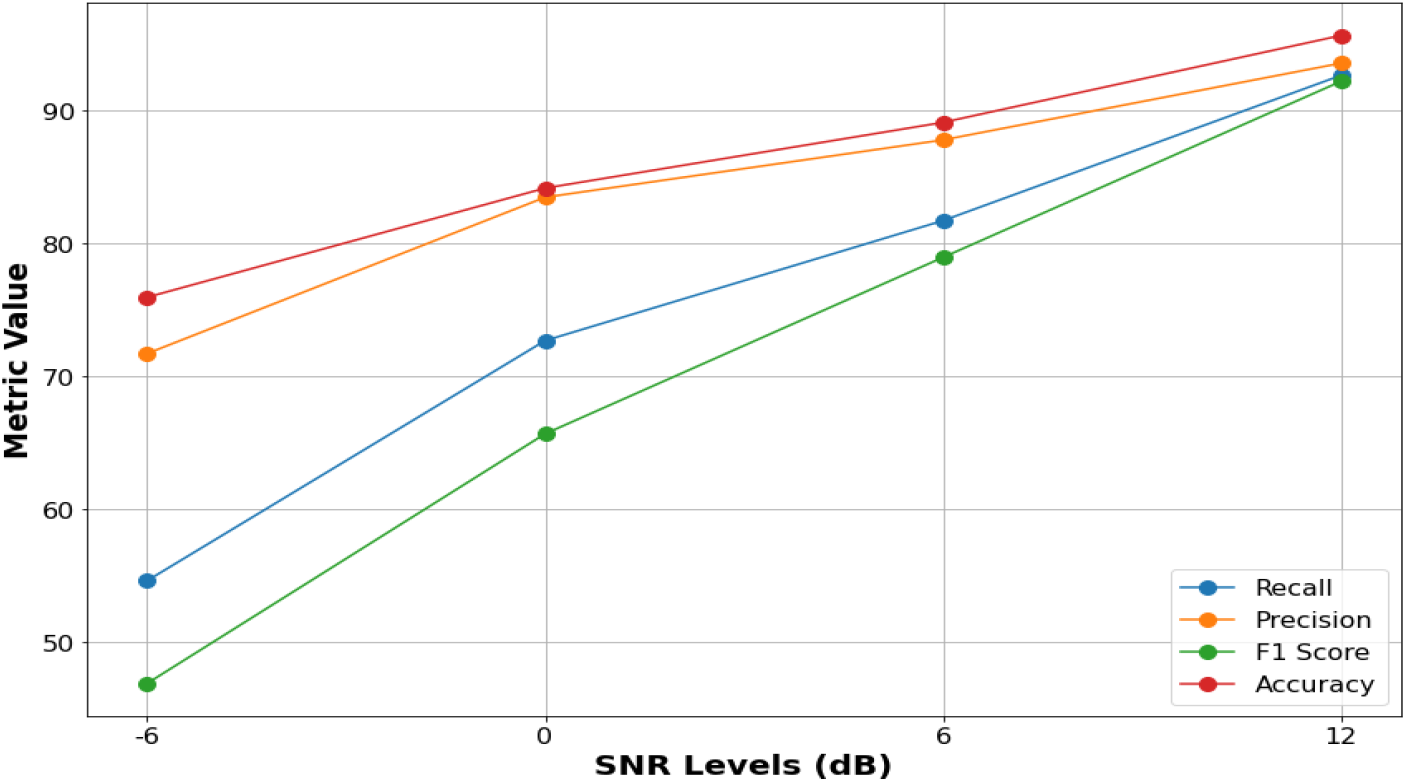
Recall, precision, F1-score, and accuracy of QDA classifier with different SNR levels.

### 3.3. Computational Efficiency

In addition to accuracy, efficient computational performance is a critical requirement for real-time biomedical applications, and the proposed method is well-suited to meeting this need. Fitting the shape model to a target beat has a direct closed-form solution and does not require complex iterative optimization methods. Determining the ***b*** values to fit the model to a target beat involves simply subtracting the mean beat from the target beat followed by performing a straightforward matrix-vector multiplication, all of which are simple mathematical operations. Furthermore, the method does not require large memory resources, as it relies on a compact 19×450 matrix and a lightweight trained classifier. Regarding the execution time, the average time to fit a heartbeat in the test set was 132 microseconds. Also, the average time to classify a heartbeat in the test set was found to be 16.33 microseconds for RF, 43.65 microseconds for SVM, 24.93 microseconds for ET, and 1.29 microseconds for QDA. The simplicity, speed, and low memory requirements of the proposed method emphasize its robustness for real-time applications, such as continuous patient monitoring and early disease detection. The ability to fit and classify heartbeats in mere microseconds demonstrates the feasibility of integrating this approach into embedded systems, which paves the way for efficient and accessible healthcare solutions.

### 3.4. Comparison With Previous Methods

The comparison between the proposed method and other approaches reported in the literature is presented in Table 5. It is evident that the proposed method with QDA classifier exhibits superior performance across all evaluation metrics. The proposed method demonstrates balanced performance across key metrics, in contrast to other approaches that tend to excel in certain metrics while underperforming in others **[39–41]**. In addition to comparison based on evaluation metrics, other crucial aspects include model efficiency, generalizability, and noise resilience. Although the model in **[42]** is resource-efficient, it relies on the detection of ECG fiducial points, which becomes challenging in noisy environments, such as those encountered in ambulatory ECG recordings. Similarly, while the model in **[43]** demonstrates high performance across all metrics, it was validated using the intra-patient scheme, which leads to inflated performance metrics and reduces the model’s ability to generalize to unseen patients. Furthermore, although the authors of **[44]** evaluated their model on the MIT-BIH Arrhythmia Database without applying any preprocessing, the predominance of high-quality ECG segments within the dataset masks the model’s limitations when handling noisier or more challenging signals. Additionally, their model is not devoid from other limitations, including computational complexity and the assumption that each ECG segment contains only one type of arrhythmia. In summary, the proposed method not only achieves superior and balanced performance across all evaluation metrics but also addresses key limitations of existing approaches, such as sensitivity to noise, overfitting to intra-patient schemes, and computational complexity, thereby demonstrating its practicality and generalizability in real-world applications.

**Table 5.** Comparison of different methods against the proposed model with SVM classifier.

| Study | Method | Heartbeat<br>Classes | Recall | Precision | Specificity | F1 Score | Accuracy |
| --- | --- | --- | --- | --- | --- | --- | --- |
| [39] | Wavelet transform<br>& CNN | PVC, non-PVC | 82.55% | 82.39% | 98.68% | 82.47% | 97.56% |
| [40] | Rules | N, PAC, PVC | 76.54% | 90.47% | --- | --- | 85.56% |
| [41] | DenseNet-GRU | N, S, V, F, Q | 82% | --- | 96% | --- | 92% |
| [42] | Multi-feature fusion<br>& bidirectional<br>LSTM | N, PB, LBBB,<br>RBBB, PVC | --- | --- | --- | 93.18% | 96.1% |
| [43] | Ensemble of CNN<br>and LSTM | N, A, LBBB,<br>RBBB, V | 98.37% | --- | 99.59% | 98.41% | 99.35% |
| [44] | LSTM and CNN | N, APB, LBBB,<br>RBBB, PVC | 97.50% | --- | 98.70% | 98.11% | 98.10% |
| Proposed<br>(without noise) | SSM Fitting +<br>SVM | N, MI, PVC,<br>RBBB | 97.49% | 97.73% | --- | 97.58% | 98.63% |
| Proposed<br>(with noise) | SSM Fitting +<br>SVM | N, MI, PVC,<br>RBBB | 95.49% | 95.57% | --- | 95.49% | 97.44% |

## 4. Conclusions

We introduce a novel method for automated classification of various heartbeat types based on statistical shape model fitting. This study is the first of its kind that utilizes SSMs for classifying ECG abnormalities. The proposed method offers several advantages, including accurate classification of several types of heartbeats with diverse morphologies, excellent noise-resistance with good accuracy even at high levels of noise, and computationally efficiency, indicating its potential use in wearable cardiac monitors. These features make the proposed method a reliable solution for automatic heartbeat classification and real-time heartbeat monitoring. Future efforts will focus on including other cardiac diseases in addition to applying the proposed method for classifying ECG beats acquired with fewer leads.

## Data Availability

The data used in this study is a subset of the PhysioNet/Computing in Cardiology Challenge 2021, which is openly accessible in [physionet] at [https://physionet.org/content/challenge-2021/1.0.3/]. Data will be made available upon request.

## Appendix A

**Table A1.** Default parameters of RF, SVM, ET, and QDA classifiers.

| Classifier |  |  |  |  |  |  |  |
| --- | --- | --- | --- | --- | --- | --- | --- |
| RF |  | SVM |  | ET |  | QDA |  |
| Parameter | Value | Parameter | Value | Parameter | Value | Parameter | Value |
| bootstrap | True | C | 1.0 | bootstrap | False | priors | None |
| ccp_alpha | 0.0 | break_ties | False | ccp_alpha | 0.0 | reg_param | 0.0 |
| class_weight | None | cache_size | 200 | class_weight | None | store_covariance | False |
| criterion | 'gini' | class_weight | None | criterion | 'gini' | tol | 0.0001 |
| max_depth | None | coef0 | 0.0 | max_depth | None | ----- | ----- |
| max_features | 'sqrt' | decision_function_shape | 'ovr' | max_features | 'sqrt' | ----- | ----- |
| max_leaf_nodes | None | degree | 3 | max_leaf_nodes | None | ----- | ----- |
| max_samples | None | gamma | 'scale' | max_samples | None | ----- | ----- |
| min_impurity_decrease | 0.0 | kernel | 'rbf' | min_impurity_decrease | 0.0 | ----- | ----- |
| min_samples_leaf | 1 | max_iter | -1 | min_samples_leaf | 1 | ----- | ----- |
| min_samples_split | 2 | probability | False | min_samples_split | 2 | ----- | ----- |
| min_weight_fraction_leaf | 0.0 | random_state | None | min_weight_fraction_leaf | 0.0 | ----- | ----- |
| n_estimators | 100 | shrinking | True | n_estimators | 100 | ----- | ----- |
| n_jobs | None | tol | 0.001 | n_jobs | None | ----- | ----- |
| oob_score | False | verbose | False | oob_score | False | ----- | ----- |
| random_state | None | ----- | ----- | random_state | None | ----- | ----- |
| verbose | 0 | ----- | ----- | verbose | 0 | ----- | ----- |
| warm_start | False | ----- | ----- | warm_start | False | ----- | ----- |

## References

1. Cardiovascular Diseases. Available online: https://www.who.int/health-topics/cardiovascular-diseases.

2. Adib, E.; Fernandez, A.S.; Afghah, F.; Prevost, J.J. Synthetic ECG Signal Generation Using Probabilistic Diffusion Models. IEEE Access 2023, 11, 75818–75828, doi:10.1109/ACCESS.2023.3296542.

3. Hong, S.; Zhou, Y.; Shang, J.; Xiao, C.; Sun, J. Opportunities and Challenges of Deep Learning Methods for Electrocardiogram Data: A Systematic Review. Computers in Biology and Medicine 2020, 122, 103801, doi:10.1016/j.compbiomed.2020.103801.

4. Abdou, A.; Krishnan, S. Enhancement of Single-Lead Dry-Electrode ECG through Wavelet Denoising. Front. Signal Process. 2024, 4, 1396077, doi:10.3389/frsip.2024.1396077.

5. Martínez-Sellés, M.; Marina-Breysse, M. Current and Future Use of Artificial Intelligence in Electrocardiography. JCDD 2023, 10, 175, doi:10.3390/jcdd10040175.

6. Sannino, G.; De Pietro, G. A Deep Learning Approach for ECG-Based Heartbeat Classification for Arrhythmia Detection. Future Generation Computer Systems 2018, 86, 446–455, doi:10.1016/j.future.2018.03.057.

7. Moody, G.B.; Mark, R.G. The Impact of the MIT-BIH Arrhythmia Database. IEEE Eng Med Biol Mag 2001, 20, 45–50, doi:10.1109/51.932724.

8. Sultan Qurraie, S.; Ghorbani Afkhami, R. ECG Arrhythmia Classification Using Time Frequency Distribution Techniques. Biomed. Eng. Lett. 2017, 7, 325–332, doi:10.1007/s13534-017-0043-2.

9. Kiranyaz, S.; Ince, T.; Gabbouj, M. Real-Time Patient-Specific ECG Classification by 1-D Convolutional Neural Networks. IEEE Trans. Biomed. Eng. 2016, 63, 664–675, doi:10.1109/TBME.2015.2468589.

10. Luz, E.J.D.S.; Nunes, T.M.; De Albuquerque, V.H.C.; Papa, J.P.; Menotti, D. ECG Arrhythmia Classification Based on Optimum-Path Forest. Expert Systems with Applications 2013, 40, 3561–3573, doi:10.1016/j.eswa.2012.12.063.

11. Acharya, U.R.; Fujita, H.; Lih, O.S.; Hagiwara, Y.; Tan, J.H.; Adam, M. Automated Detection of Arrhythmias Using Different Intervals of Tachycardia ECG Segments with Convolutional Neural Network. Information Sciences 2017, 405, 81–90, doi:10.1016/j.ins.2017.04.012.

12. Moody, G.B.; Mark, R.G. MIT-BIH Atrial Fibrillation Database 1992. doi: 10.13026/C2MW2D

13. Nolle, F.M.; Bowser, R.W. Creighton University Ventricular Tachyarrhythmia Database 1992. doi:10.13026/C2X59M

14. Teijeiro, T.; Felix, P.; Presedo, J.; Castro, D. Heartbeat Classification Using Abstract Features From the Abductive Interpretation of the ECG. IEEE J. Biomed. Health Inform. 2018, 22, 409–420, doi:10.1109/JBHI.2016.2631247.

15. Gupta, A.; Banerjee, A.; Babaria, D.; Lotlikar, K.; Raut, H. Prediction and Classification of Cardiac Arrhythmia. In Proceedings of the Sentimental Analysis and Deep Learning; Shakya, S., Balas, V.E., Kamolphiwong, S., Du, K.-L., Eds.; Springer Singapore: Singapore, 2022; Vol. 1408, pp. 527–538. doi: 10.1007/978-981-16-5157-1_41

16. H. Guvenir, B. Acar, Arrhythmia 1997. doi: 10.24432/C5BS32

17. Xiao, Q.; Lee, K.; Mokhtar, S.A.; Ismail, I.; Pauzi, A.L.B.M.; Zhang, Q.; Lim, P.Y. Deep Learning-Based ECG Arrhythmia Classification: A Systematic Review. Applied Sciences 2023, 13, 4964, doi:10.3390/app13084964.

18. Di Paolo, í.F.; Castro, A.R.G. Intra- and Interpatient ECG Heartbeat Classification Based on Multimodal Convolutional Neural Networks with an Adaptive Attention Mechanism. Applied Sciences 2024, 14, 9307, doi:10.3390/app14209307.

19. Jeong, Y.; Lee, J.; Shin, M. Enhancing Inter-Patient Performance for Arrhythmia Classification with Adversarial Learning Using Beat-Score Maps. Applied Sciences 2024, 14, 7227, doi:10.3390/app14167227.

20. Statistical Shape and Deformation Analysis: Methods, Implementation and Applications; Zheng, G., Li, S., Székely, G., Eds.; Computer vision and pattern recognition series; Academic Press, an imprint of Elsevier: London, 2017; ISBN 978-0-12-810493-4.

21. Cootes, T.F.; Taylor, C.J. Statistical Models of Appearance for Medical Image Analysis and Computer Vision.; Sonka, M., Hanson, K.M., Eds.; San Diego, CA, July 3 2001; pp. 236–248. doi: 10.1117/12.431093

22. Cootes, T. An Introduction to Active Shape Models. 2000. Available online:

23. Ambellan, F.; Lamecker, H.; Von Tycowicz, C.; Zachow, S. Statistical Shape Models: Understanding and Mastering Variation in Anatomy. In Biomedical Visualisation; Rea, P.M., Ed.; Advances in Experimental Medicine and Biology; Springer International Publishing: Cham, 2019; Vol. 1156, pp. 67–84 ISBN 978-3-030-19384-3.

24. Su, Z. Statistical Shape Modelling: Automatic Shape Model Building. PhD, University College London, 2011.

25. Premature Ventricular Contractions (Premature Ventricular Complex, Premature Ventricular Beats). Available online: https://ecgwaves.com/topic/premature-ventricular-contractions-complex-beats-ecg/.

26. Salari, N.; Morddarvanjoghi, F.; Abdolmaleki, A.; Rasoulpoor, S.; Khaleghi, A.A.; Hezarkhani, L.A.; Shohaimi, S.; Mohammadi, M. The Global Prevalence of Myocardial Infarction: A Systematic Review and Meta-Analysis. BMC Cardiovasc Disord 2023, 23, 206, doi:10.1186/s12872-023-03231-w.

27. Right Bundle Branch Block (RBBB): ECG, Criteria, Definitions, Causes & Treatment. Available online: https://ecgwaves.com/topic/right-bundle-branch-block-rbbb-ecg-criteria-treatment/.

28. Reyna, M.; Sadr, N.; Gu, A.; Perez Alday, E.A.; Liu, C.; Seyedi, S.; Shah, A.; Clifford, G. Will Two Do? Varying Dimensions in Electrocardiography: The PhysioNet/Computing in Cardiology Challenge 2021. doi: 10.13026/gt86-a263.

29. Goldberger, A.L.; Amaral, L.A.; Glass, L.; Hausdorff, J.M.; Ivanov, P.C.; Mark, R.G.; Mietus, J.E.; Moody, G.B.; Peng, C.K.; Stanley, H.E. PhysioBank, PhysioToolkit, and PhysioNet: Components of a New Research Resource for Complex Physiologic Signals. Circulation 2000, 101, E215–220, doi: 10.1161/01.cir.101.23.e215.

30. Liu, F.; Liu, C.; Zhao, L.; Zhang, X.; Wu, X.; Xu, X.; Liu, Y.; Ma, C.; Wei, S.; He, Z.; et al. An Open Access Database for Evaluating the Algorithms of Electrocardiogram Rhythm and Morphology Abnormality Detection. j med imaging hlth inform 2018, 8, 1368–1373, doi: 10.1166/jmihi.2018.2442.

31. Wagner, P.; Strodthoff, N.; Bousseljot, R.-D.; Samek, W.; Schaeffter, T. PTB-XL, a Large Publicly Available Electrocardiography Dataset. doi: 10.13026/kfzx-aw45.

32. Georgia 12-Lead ECG Challenge Database. Available online: https://www.kaggle.com/datasets/bjoernjostein/georgia-12lead-ecg-challenge-database (accessed on 16 November 2024).

33. Zheng, J.; Zhang, J.; Danioko, S.; Yao, H.; Guo, H.; Rakovski, C. A 12-Lead Electrocardiogram Database for Arrhythmia Research Covering More than 10,000 Patients. Sci Data 2020, 7, 48, doi:10.1038/s41597-020-0386-x.

34. Zheng, J.; Chu, H.; Struppa, D.; Zhang, J.; Yacoub, S.M.; El-Askary, H.; Chang, A.; Ehwerhemuepha, L.; Abudayyeh, I.; Barrett, A.; et al. Optimal Multi-Stage Arrhythmia Classification Approach. Sci Rep 2020, 10, 2898, doi:10.1038/s41598-020-59821-7.

35. Chapman-Shaoxing 12-Lead ECG Database. Available online: https://www.kaggle.com/datasets/erarayamorenzomuten/chapmanshaoxing-12lead-ecg-database.

36. Elsahmarany, L.; Alshammari, M.; Tamal, M.; Alomari, A. A Robust Continuous Wavelet Transform (CWT) Based for R-Peak Detection Method of ECG 2023. doi: 10.1101/2023.07.31.23293050.

37. Alhalabi, A.; Alzahrani, S.; Tamal, M. Robust and Parameter Free QRS Complex Detection in ECG for Heart Disease Diagnosis and Monitoring. Unpublished Work.

38. Pedregosa, F.; Varoquaux, G.; Gramfort, A.; Michel, V.; Thirion, B.; Grisel, O.; Blondel, M.; Prettenhofer, P.; Weiss, R.; Dubourg, V.; et al. Scikit-Learn: Machine Learning in Python. MACHINE LEARNING IN PYTHON.

39. Li, Q.; Liu, C.; Li, Q.; Shashikumar, S.P.; Nemati, S.; Shen, Z.; Clifford, G.D. Ventricular Ectopic Beat Detection Using a Wavelet Transform and a Convolutional Neural Network. Physiol. Meas. 2019, 40, 055002, doi:10.1088/1361-6579/ab17f0.

40. Cai, Z.; Li, J.; Johnson, A.E.W.; Zhang, X.; Shen, Q.; Zhang, J.; Liu, C. Rule-Based Rough-Refined Two-Step-Procedure for Real-Time Premature Beat Detection in Single-Lead ECG. Physiol. Meas. 2020, 41, 054004, doi:10.1088/1361-6579/ab87b4.

41. Guo, L.; Sim, G.; Matuszewski, B. Inter-Patient ECG Classification with Convolutional and Recurrent Neural Networks. Biocybernetics and Biomedical Engineering 2019, 39, 868–879, doi:10.1016/j.bbe.2019.06.001.

42. Nikandish, R.; He, J.; Haghi, B. Resource-Efficient Heartbeat Classification Using Multi-Feature Fusion and Bidirectional LSTM 2024. doi: 10.48550/arXiv.2405.15312.

43. Irfan, S.; Anjum, N.; Althobaiti, T.; Alotaibi, A.A.; Siddiqui, A.B.; Ramzan, N. Heartbeat Classification and Arrhythmia Detection Using a Multi-Model Deep-Learning Technique. Sensors 2022, 22, 5606, doi:10.3390/s22155606.

44. Oh, S.L.; Ng, E.Y.K.; Tan, R.S.; Acharya, U.R. Automated Diagnosis of Arrhythmia Using Combination of CNN and LSTM Techniques with Variable Length Heart Beats. Computers in Biology and Medicine 2018, 102, 278–287, doi:10.1016/j.compbiomed.2018.06.002.

